# Quantifying the importance and location of SARS-CoV-2 transmission events in large metropolitan areas

**DOI:** 10.1101/2020.12.15.20248273

**Authors:** Alberto Aleta, David Martín-Corral, Michiel A. Bakker, Ana Pastore y Piontti, Marco Ajelli, Maria Litvinova, Matteo Chinazzi, Natalie E. Dean, M. Elizabeth Halloran, Ira M. Longini, Alex Pentland, Alessandro Vespignani, Yamir Moreno, Esteban Moro

## Abstract

Detailed characterization of SARS-CoV-2 transmission across different settings can help design less disruptive interventions. We used real-time, privacy-enhanced mobility data in the New York City and Seattle metropolitan areas to build a detailed agent-based model of SARS-CoV-2 infection to estimate the where, when, and magnitude of transmission events during the pandemic’s first wave. We estimate that only 18% of individuals produce most infections (80%), with about 10% of events that can be considered super-spreading events (SSEs). Although mass-gatherings present an important risk for SSEs, we estimate that the bulk of transmission occurred in smaller events in settings like workplaces, grocery stores, or food venues. The places most important for transmission change during the pandemic and are different across cities, signaling the large underlying behavioral component underneath them. Our modeling complements case studies and epidemiological data and indicates that real-time tracking of transmission events could help evaluate and define targeted mitigation policies.

## Introduction

Without effective pharmaceutical interventions, the COVID-19 pandemic triggered the implementation of severe mobility restrictions and social distancing measures worldwide aimed at slowing down the transmission of SARS-CoV-2. From shelter in place orders to closing restaurants/shops or restricting travel, the rationale of those measures is to reduce the number of social contacts, thus breaking transmission chains. Though individuals may remain highly connected to household members or close contacts, these measures reduce the connections in the general community that allow the virus to move through the network of human contacts. Some venues may attract more individuals from otherwise unconnected social networks, or may attract individuals who are more active and thus have greater exposure. Understanding how interventions targeted at particular venues could impact transmission of SARS-CoV-2 can help us devise better non-pharmaceutical interventions (NPIs) that pursue public health objectives while minimizing disruption to the economy, the education system, and other facets of everyday life.

Although it is by now clear that NPIs have helped to mitigate the COVID-19 pandemic^1^, most of the evidence is based on measuring the subsequent reduction in the case growth rate or secondary reproductive number. For example, econometric models were used to estimate the effect of the introduction of NPIs on the secondary reproductive number^2, 3^. Other studies have shown directly (through correlations or statistical models^4^) or indirectly (through epidemic simulations^5, 6^) the relationship between mobility or individuals’ activity and number of cases. Unfortunately, most of the data used so far do not have the granularity required to assess how social contacts and SARS-CoV-2 transmission events are modified by NPIs^7^.

This is especially important given the heterogeneous spreading of SARS-CoV-2. Overdispersion in the number of secondary infections produced by a single individual was an important characteristic of the 2003 SARS pandemic^8^ and has been similarly observed for SARS-CoV-2^9^. Several drivers of super-spreading events (SSEs) have been proposed: biological, due to differences in individuals’ infectiousness; behavioral, caused by unusually large gatherings of contacts; and environmental, in places where the surrounding conditions facilitate spread^10^. Transmissibility depends critically on the characteristics of the place where contacts happen, with many SSEs documented in crowded, indoor events with poor ventilation. A characteristic of this overdispersion is that most infections (around 80%) are due to a small number of people or places (20%), suggesting that better targeted NPIs or cluster-based contact tracing strategies can be devised to control the pandemic^11^. Although several studies have provided insights on SSEs^7, 12^, given their outsized importance for SARS-CoV-2, we need better information about where, when, and to what extent these SSEs happen and how they may be mitigated or amplified by NPIs.

In this paper we use a longitudinal database of detailed mobility and socio-demographic data to estimate the probability of contact and transmission between individuals in different places across the New York and Seattle metropolitan areas, during the period from February 17 to June 1 of 2020 (see Supp. Material Section 1). Note that the metropolitan areas considered extend beyond the city limits for both locations. We selected these areas because of their large differences in COVID-19 epidemiology, population size and density. The NY metro area has a population of 20 million people, while the Seattle metro area has 3.8 million inhabitants. Moreover, the NY metro area has a higher density (5,438 people per km^2^, median by census tract) than Seattle (1,576 people per km^2^). Finally the number of reported COVID-19 cases/deaths during the study period in the NY area was very large (223 per 100,000) compared to that in the Seattle area (24 per 100,000). Individual mobility data is sampled to be representative of the different census areas (Census Block Groups, see Figure 1). Probabilistic estimation of contact between individuals is weighted according to the likelihood of exposure between them in the different places around the metro areas. This defines a weighted temporal network consisting of four layers representing the probabilistic estimation of physical/social interactions occurring in (1) the community, (2) workplaces, (3) households, and (4) schools, see Figure 1. The community and workplaces layers are generated using 4 months of data observed in the New York and Seattle metropolitan areas from anonymized users who opted-in to provide access to their location data, through a GDPR-compliant (General Data Protection Regulation) framework provided by Cuebiq (see Supp. Material Section 1).

**Figure 1.**
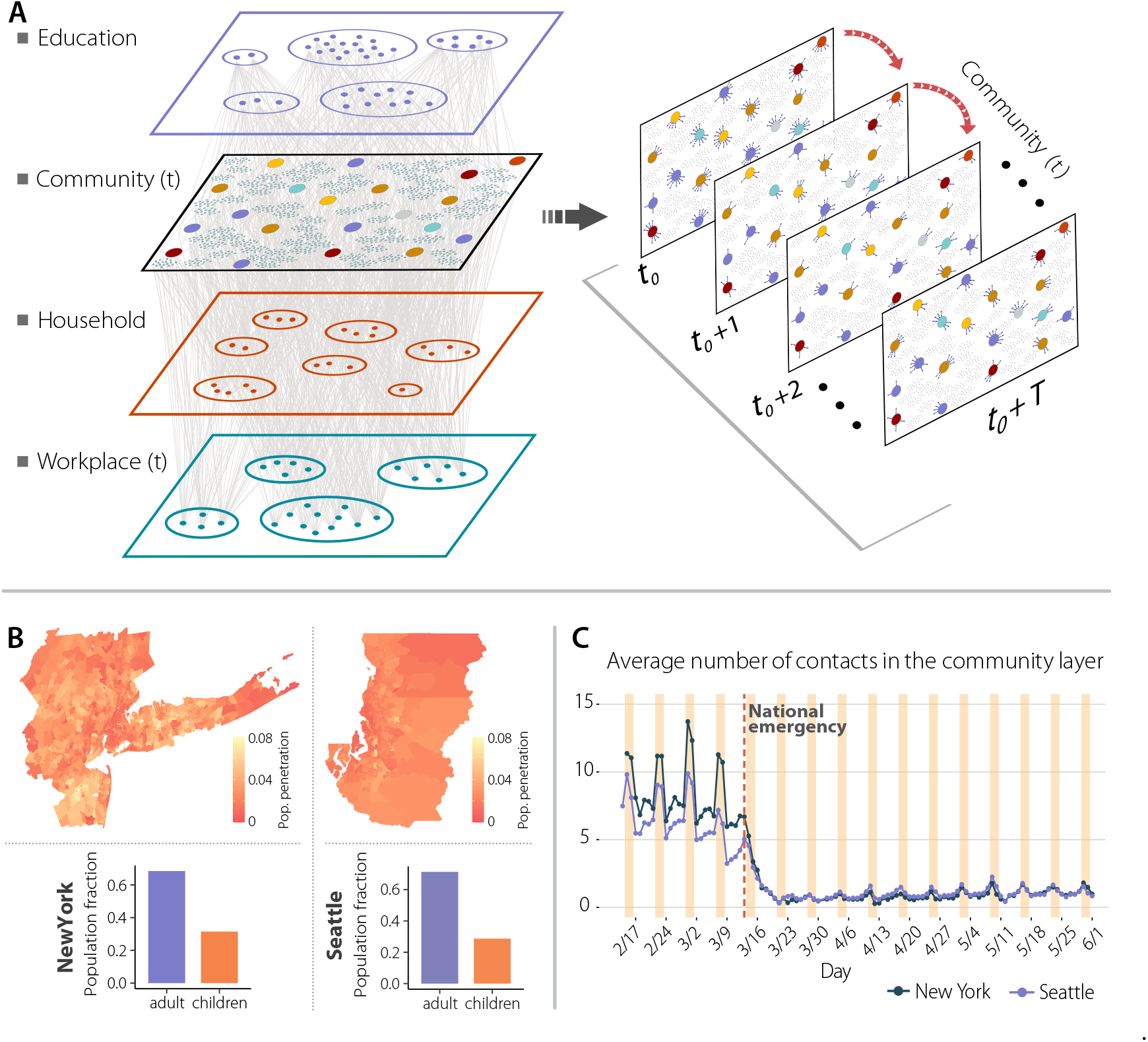
Network components, New York and Seattle metropolitan areas population and social contacts dynamics at the community layer over time. Panel **a** is a schematic illustration of the weighted multilayer and temporal network for our synthetic population built from mobility data. There are four different layers; the school and household layers are static over time, and the combined workplace and community layers have a daily temporal component. Panel **b** shows the geographic penetration (fraction of mobile devices by population) from our mobility data compared to the total population for the New York and Seattle metropolitan areas. Panel **c** represents the average daily number of contacts in the community layer for both metropolitan areas.

The data allows us to understand how infection can propagate in each layer by estimating the probability of transmission between individuals in the same setting, including schools, workplaces, households and multiple locations in the community. Settings associated to the community are obtained from a large database of 375k locations in the New York and 70k in the Seattle from the Foursquare public API. By measuring the probability that people interact in the different layers, we construct a probabilistic time-varying contact network of *ω*_*i jt*_ between individuals *i* and *j* on the same day *t* in the education, community, work and household layers. Estimates of transmission in the community layer is done by extracting stays of users to the settings using different time and distance in the setting. Our results are independent of the particular choice of minimal time (5 minutes or 15min) and maximum distance to the setting (10 meters or 50 meters), see Figure 1 and Supp. Material Section 1 and 2 for more information about the data and layers. Our model covers all possible interactions in urban areas and not just foot traffic to commercial locations that people visit^7^, something especially important given the relevant role of households, schools or workplaces in the transmission of the SARS-CoV-2. It is important to note that the underlying data does not provide a direct measurement of contacts between individuals and the nature of these contacts (masked/unmasked, with conversation). Rather, our method uses this data to extrapolate the locations visited by each subject and the amount of time they spent there, in order to estimate the transmission probability between individuals, relaxing the homogeneous mixing assumption commonly used in mathematical modeling approaches. In simpler terms, our method does not detect directly co-location of individuals, but rather is a probabilistic estimation of the transmission between them according to the time they spend in the same places or layers.

To model the natural history of the SARS-CoV-2 infection, we implemented a stochastic, discrete-time compartmental model on top of the contact network *ω*_*i jt*_ in which individuals transition from one state to the other according to the distributions of key time-to-event intervals (e.g., incubation period, serial interval, etc.) as per available data on SARS-CoV-2 transmission (see Supp. Material Section 3 for details). In the infection transmission model, susceptible individuals (S) become infected through contact with any of the infectious categories (infectious symptomatic (IS), infectious asymptomatic (IA) and pre-symptomatic (PS)), transitioning to the latent compartment (L), where they are infected but not infectious yet. Latent individuals branch out in two paths according to whether the infection will be symptomatic or not. We also consider that symptomatic individuals experience a pre-symptomatic phase and that once they develop symptoms, they can experience diverse degrees of illness severity, leading to recovery (R) or death (D). The value of the basic reproduction number is calibrated to the weekly number of deaths (see Supp. Material Sections 4, 5 and 7 for further information on the calibration process, model’s details, and for the sensitivity of our results towards different values of parameters used in the model).

## Results

### Impact of NPIs

Our data clearly show that the statistic of potential contacts in the two metro areas have changed due to the introduction of NPIs during the week of March 15th to March 22nd, see Figure 1. A National Emergency was declared on March 13th, and the NY City School System announced the closure of schools in March 16th^13^. NY City Mayor issued a “shelter in place” order in the city on March 17^14^, and non-essential business were ordered to close or suspend all in-person functions in New York, New Jersey and Connecticut by March 22nd. As we can see in Figure 1 the individuals’ total number of contacts decreased dramatically from around 7 (in our community layer) to below 2. In Seattle, the reduction of contacts started one week earlier than in NY City, coinciding with earlier closing of some schools^15^, and the Seattle mayor issuing a proclamation of civil emergency on March 3rd^16^.

In Figure 2 we report numerical simulations of the epidemic curve that accurately reproduce the evolution of the incidence of new COVID-19-related deaths in both NY and Seattle metro areas, even though both cities were affected very differently by the epidemic in the first wave. The analysis identifies the impact of the reduction in the estimated number of contacts due to the implemented NPIs: both in the NY and Seattle metro areas, *R*_*t*_ dropped below 1 one week after NPIs were introduced. To estimate the importance of timely implementations of NPIs in metropolitan areas, we have generated counterfactual scenarios in which the NPIs and the ensuing reduction in the number of contacts could have happened one week earlier or later than the actual timeline^19^. The comparison between NY and Seattle is relevant, because we observed that the reduction in contacts in Seattle started to happen exactly one week before that in NY. To this end we have shifted in time the contact patterns around the week where NPIs where introduced in both cities. The results for these scenarios are reported in Figure 2d, where we see that a one week delay in introducing NPIs could have yielded a peak in the number of deaths two times larger than the observed one (0.7 deaths per 1,000 people compared to the 0.35 per 1,000). This doubling in peak deaths following a one week delay is also observed in the Seattle metro area and in the cumulative infection prevalence in the metro area. Conversely, a one week earlier implementation of the NPIs timeline in NY area could have reduced the death peak by more than a factor of three, a result similar to that found using county-level simulations^19^. In Seattle, implementing the NPIs one week earlier would have prevented the first wave of infections. For this reason, the results are not contained in panel F.

**Figure 2.**
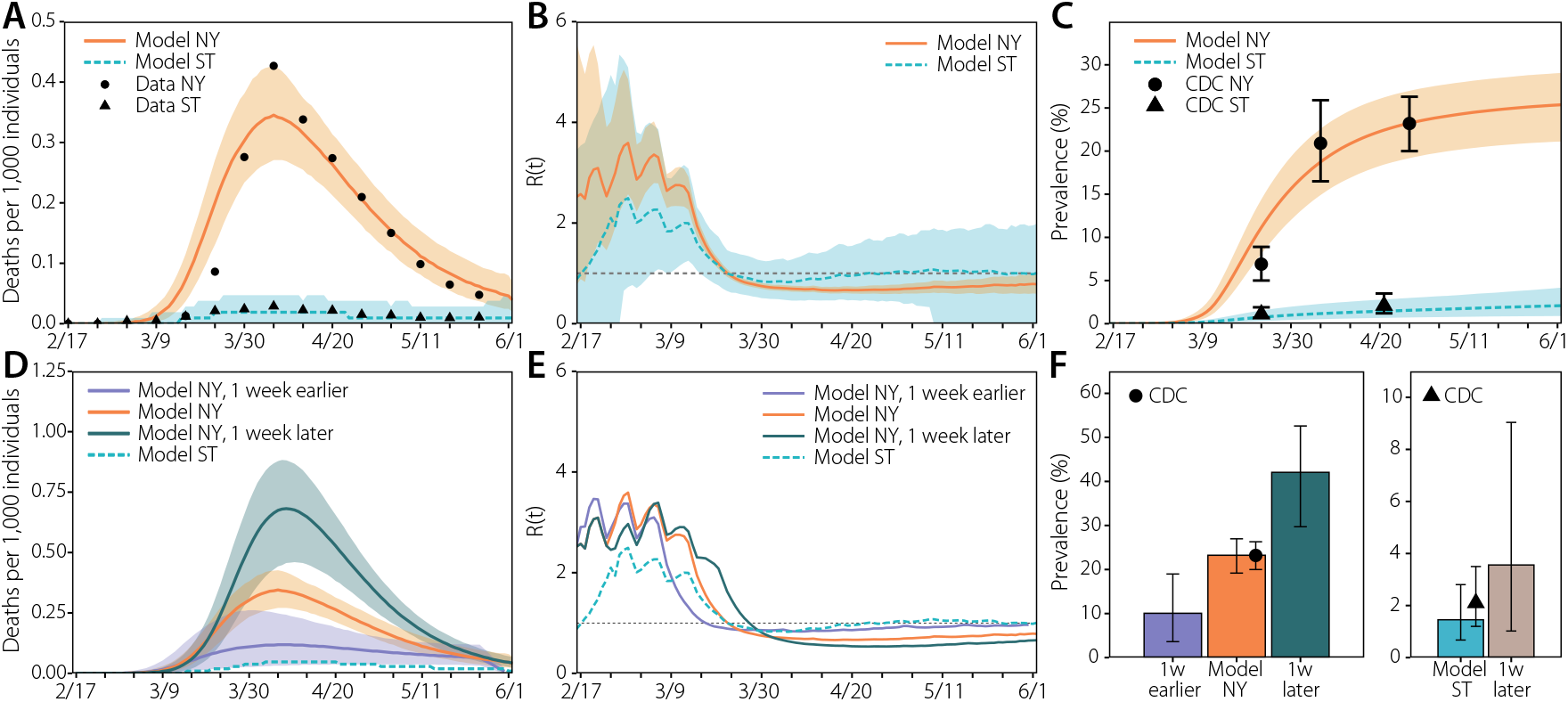
Evolution of the first wave. (a) Weekly number of deaths in New York (NY) and Seattle (ST) metro areas. The dots/triangles represent the reported surveillance data used in the calibration of the models. The lines represent the median of the model ensemble for each location and the shaded areas the 95% C.I. of the calibrated model^17^. (b) Evolution of the effective reproduction number according to the output of the simulation. The solid (dashed) line represents the median of the model ensemble and the shaded areas the 95% C.I. of the model. (c) Estimated prevalence in our model (median represented with solid/dashed lines and 95% C.I with the shaded area) and values reported by the CDC (dots/triangles represent New York and Seattle data respectively)^18^. (d) Estimated number of deaths if the NPIs had been applied in New York one week earlier/later. Solid (dashed) lines represent the median of the model ensemble and the shaded areas the 95% C.I. (e) Estimated evolution of the effective reproduction number if the measures had been applied in New York one week earlier/later. Solid (dashed) lines represent the median of the model ensemble. (f) Estimated prevalence in New York (left) and Seattle (right) if the NPIs had been applied in New York one week earlier/later and in Seattle one week later. The height of the bars represents the median of the model ensemble, while the vertical error bars represent the 95% C.I. The dot/triangle shows the value reported by the CDC for the last week of April 2020.

### Taxonomy of transmission events

The high resolution of our dataset allows us to estimate the relevance of different settings and the effects of NPIs on the transmission dynamic of SARS-CoV-2. People spent different time in each layer and place before and after the introduction of NPIs (see Supp. Material Section 1). As a result, the number of infections varied significantly during the observed period. As we can see in Figure 3, before NPIs were introduced, we estimate that most infections took place in the community and workplace layers. Once restrictions were implemented in both cities on March 16th, as expected, the proportion of infections in the household layer greatly increased, especially in the NY area. In Seattle, the number of infections in the workplace and household layers were comparable, probably because the number of cases overall was lower than in NY. We can further stratify data by venue type in the community layer as in Figure 3, by looking at the estimated top categories (see Supp. Material Section 1 for their definition) in terms of the number of total infections throughout the whole period. Before the NPIs were introduced, our model estimates that most of the infections in the community layer happened in Food/Beverage, Shopping, and Exercise venues. Also a significant number of infections happened in Art/Museums and Sport/Events venues. After the introduction of NPIs, the number of infections in Exercise, Sport/Events or Art/Museums venues decreases as expected. However, Food, Groceries and Shopping venues became the main community setting for transmission in both cities.

**Figure 3.**
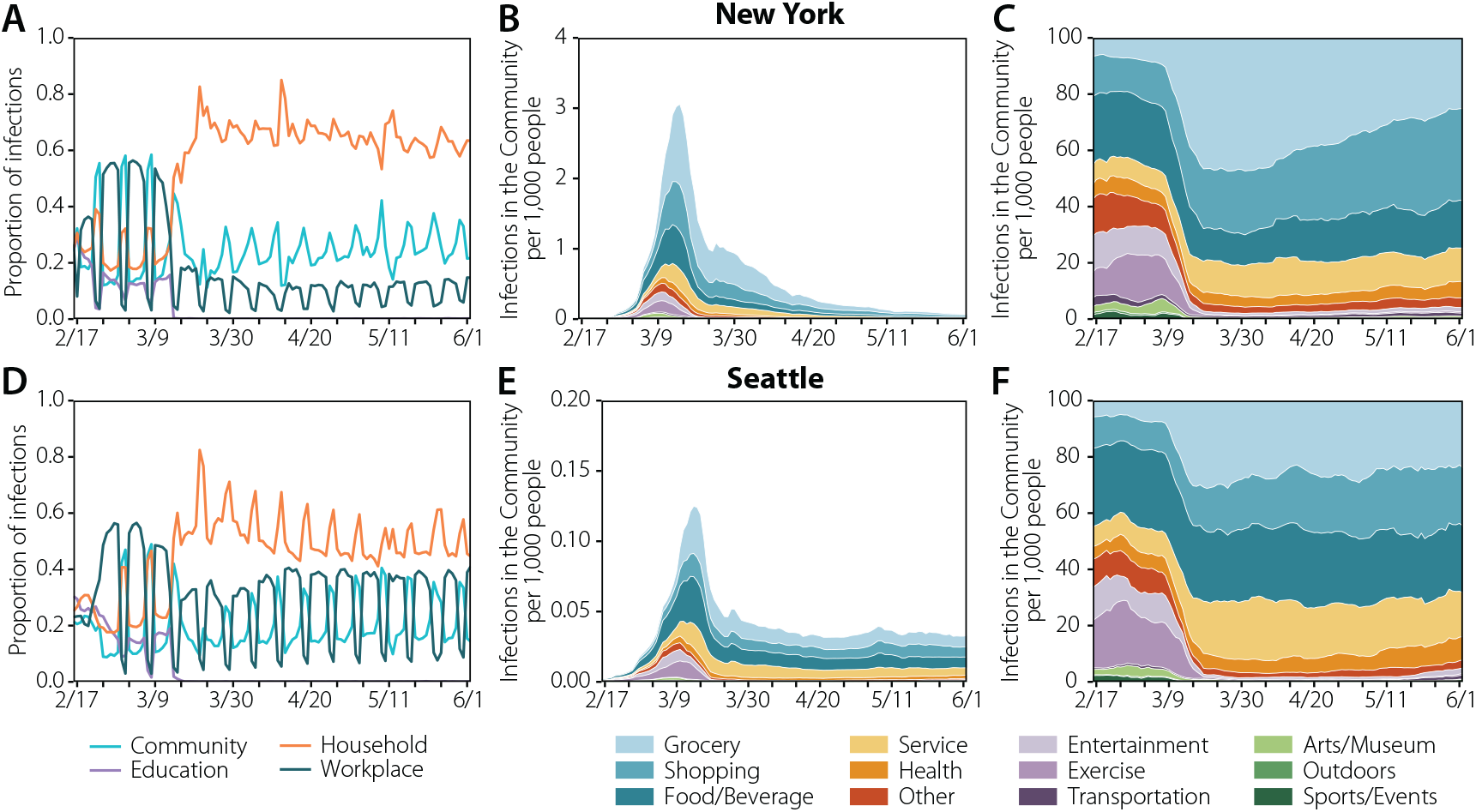
Spatial spreading of the disease. The plots in the left column represent the share of infections across layers in New York (a) and Seattle (d). In the middle column, the estimated location where the infections took place for New York (b) and Seattle (e) in the community layer. Note that the y-axis is 20 times smaller in Seattle. The evolution has been smoothed using a rolling average of 7 days. In the right column, the distributions are normalized over the total number of daily infections, showing how infections were shared across categories in the community layer. The evolution has been smoothed using a rolling average of 7 days.

### Super-spreading events

Our agent-based simulations also allow us to estimate statistically the transmission events by a single individual and estimate how many secondary infections she generates. In Figure 4 we report the distribution of the number of secondary infections produced by each individual in the community layer only. This is driven by individual-level differences in activity and those individuals they might interact with. The distribution is highly skewed and can be modeled by a negative binomial distribution with dispersion parameters (*k*) of 0.16 (NY) and 0.23 (Seattle), in agreement with the evidence accumulated from SARS-CoV-2 transmission data^9, 10, 20, 21^. As a result, super-spreading events (SSEs) are likely to be observed. We define a transmission event as a SSE if the individual infects in a specific location category more than the 99-th percentile of a Poisson distribution with average equal to *R* (see^8^ and Supp. Material Section 6 for further details), here corresponding to an infected individual infecting 8 or more others. Interestingly, if we compare the distribution of secondary infections produced before and after the introduction of NPIs, even though we see a clear reduction of SSEs, we still find a heterogeneous distribution of secondary infections. Thus, the NPIs did not prevent the formation of SSEs, but only significantly lowered their frequency.

**Figure 4.**
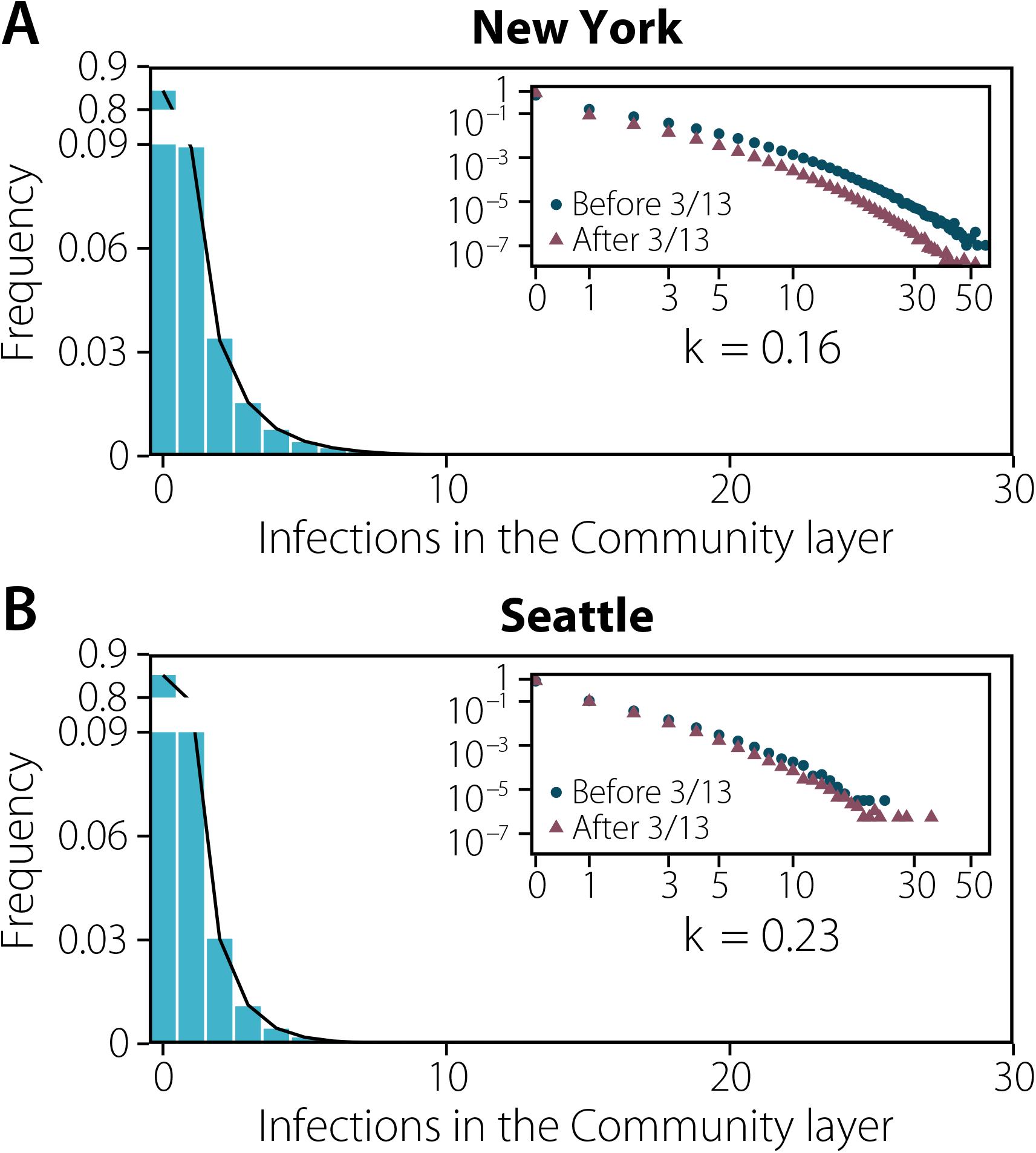
Behavioral super-spreading events. Distribution of the number of infections produced by each individual in New York (a) and Seattle (b) up to the declaration of National Emergency. The distribution is fitted to a negative binomial distribution yielding a dispersion parameter of *k* = 0.163 [0.159−0.168] 95%CI and *k* = 0.232 [0.224−0.241] 95%CI, respectively. In both plots the inset represents the same distribution on the log-scale and distinguishing infections that took place before the declaration of National Emergency on 03/13 and after that date.

Consistent with this pattern of over-dispersion in the number of transmission events, we find that the majority of infections is produced by a minority of infected people: ∼20% of infected people were responsible for more than ∼85% of the infections in both metro areas (see Figure S9 in Supp. Material). However, note that a critical driver here of this phenomenon is that a large majority of infected people (85% in the community layer) do not infect any others in our simulations. Only a small fraction of infection events (0.08%) are made of 8 (or more) secondary infections.

Transmission events and SSEs did not happen equally in different settings or along time or geography. In Figure 5 we show the results of our simulations for the total number of infections produced in each category and the share of those infections that can be related to SSEs (see also Table S2 in the Supp. Material). The combination of those two features define a continuous risk map in which places can be at different types of risk: (i) low contribution from SSEs and low contribution to the overall infections, such as Outdoor places; (ii) larger contribution from SSEs but low contribution to the overall infections, such as Sports/Events, Arts/Museum or Entertainment before the introduction of NPIs; (iii) large contribution to the overall infections but with low contribution from SSEs, such as Shopping or Food/Beverage after the introduction of NPIs; and (iv) large number of infections and with large contribution from SSEs, such as Grocery. This classification has important implications from a public health perspective. For instance, venues in (ii) do not have a major contribution to the overall infections but might represent a challenge for contact tracing. Conversely, for categories in (iii) it might be easier to trace chains of transmission but their total contribution is large. Note that this definition is not static, but changes over time due to the NPIs imposed by authorities. Indeed, looking at the weekly pattern of infections (see Fig. 5) we observe how some categories move to a different quadrant due to the behavior of individuals. Although we estimate that SSEs and infections were more likely in Arts/Museum, Sport/Events in NY, and Entertainment and Grocery in both cities, our simulations show that Grocery category still greatly contributes to the total number of infections, but do not have as many SSEs after March 16. On the other hand, we estimate that SSEs were rare before March 9 in Seattle, but their contribution doubled in the week of March 9-15 - when many individuals probably went for supplies amid preparation for the future introduction of NPIs. This observation includes implicitly a very important message: a place may not be inherently dangerous; rather, the risk is a combination of both the characteristics of the place/setting and of the behavior of individuals who visit it. This suggests revisiting studies which find that settings could play always the same role in the evolution of the pandemic^7^.

**Figure 5.**
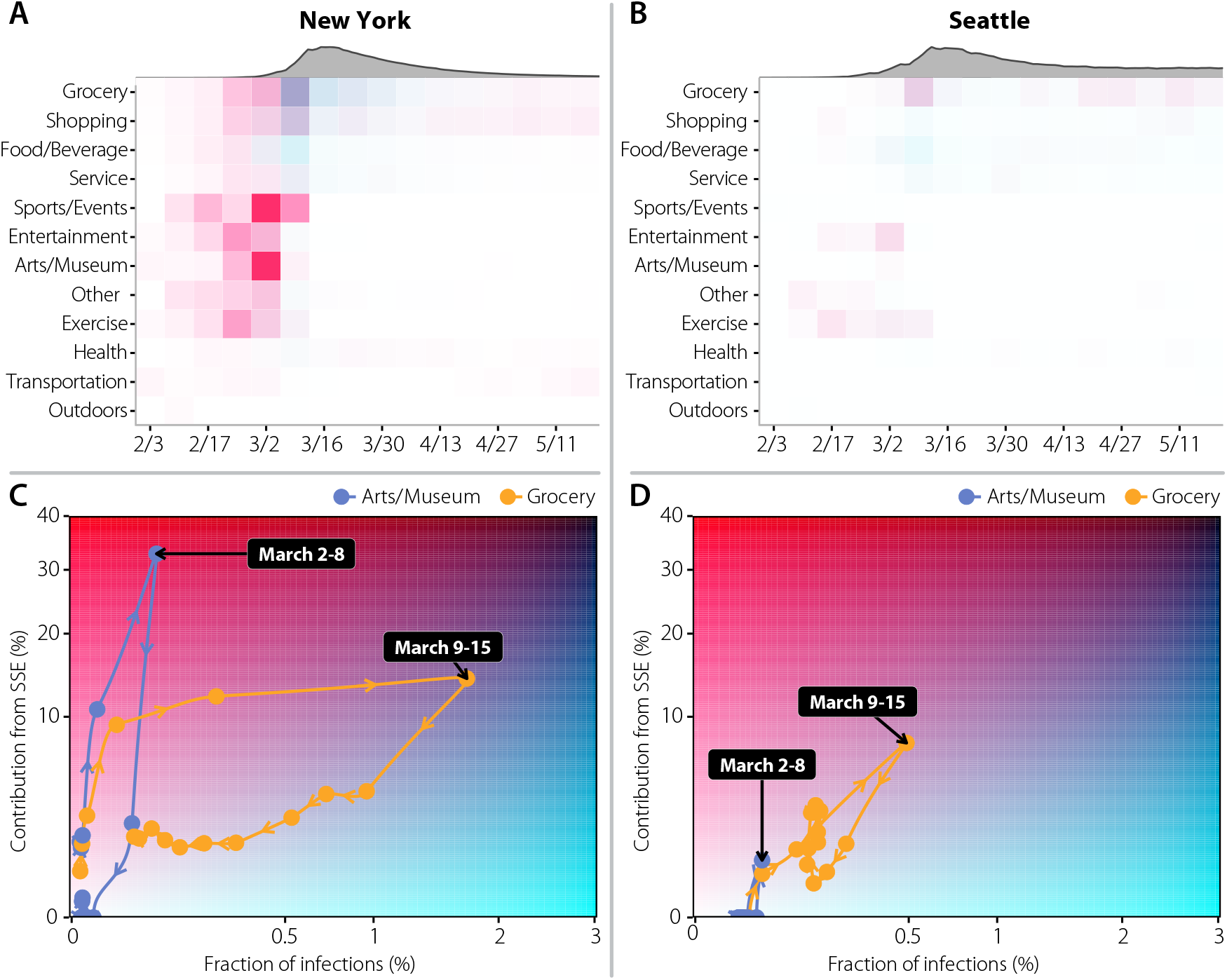
Dynamics of super-spreading events (SSE). Risk evolves with time as a function of the behavior of the population and policies in place. A) and B) : risk posed by each category per week, defined using the corresponding map below. As a reference, the gray area on top shows the estimated weekly incidence. C) and D) : the *x* axis represents the fraction of total infections that are associated with each category, while the *y* axis accounts for the share of those infections that can be attributed to SSEs in each category. Note that the fraction of infections is normalized over all the infections produced in all the social settings throughout the whole period. This defines a continuous risk map in which places with few infections and low contribution from SSEs will be situated on the left bottom corner. Places where the number of infections is high but the contribution from SSEs is low are situated in the bottom right corner. Conversely, places with large contribution from SSEs but a low amount of infections are situated on the top left corner. Lastly, places with both large number of infections and an important contribution from SSEs are situated in the top right corner. The color associated to each tile in the top row is extracted from the position of the point in the plane defined in the bottom figure. The points in the bottom row show the evolution of the position of the categories Arts/Museum and Grocery for each week, with the arrows indicating the time evolution.

## Discussion

Our results emphasize the intertwined nature of human behavior, NPIs, and the evolution of the COVID-19 pandemic in two major metropolitan areas. Specifically, our results suggest that heterogeneous connectivity and behavioral patterns among individuals lead naturally to differences in risk across settings and the generation of SSEs. In particular, the implemented partial or full closures of different settings (e.g., sport venues, museums, workplaces) had a dramatic effect in shaping the mixing patterns of the individuals outside the household^22, 23^. As a consequence, the settings responsible for the majority of transmission events and SSEs varied over time. In absolute terms, the food and beverage setting is estimated to have played a key role both in determining the number of transmission events and SSEs in the early epidemic phase; however, this setting was among the first targets of interventions and thus its contribution become zero over time because of the introduced NPIs. On the other hand, settings such as grocery stores, which consistently provided a low absolute contribution to the overall transmission and SSEs, became, in relative terms, a source of SSEs during the lockdown when most of other activities were simply not available. These findings suggest that there is room for optimizing targeted measures such as extending working time to dilute the number of contacts or the use of smart working aimed at reducing the chance of SSEs. That could be especially relevant to avoid local flare ups of cases when the reproduction number is slightly above or below the epidemic threshold.

Although the overall picture emerging from studying Seattle and New York is consistent, it is important to stress that each urban area might have specific peculiarities due to local transportation, tourism, or other economic drivers differentiating the cities’ life cycle. Our results suggest that a one-size-fits-all solution to minimize the spread of SARS-CoV-2 might have very different impact across cities. Furthermore, the results presented may not be generalized to rural areas. Though large parts of the Seattle metro area could be considered as rural, individual connectivity patterns may be differently constrained by the generally lower population density in some other parts of the country.

Our modeling analysis does not have the ambition to substitute field investigations, which remain the primary source of evidence. Some of the reported findings (e.g., the role of food and beverage venues or groceries) appear to be in agreement with epidemiological investigations^7, 24–27^. Future empirical analyses could provide further validation of our findings. Our modeling investigation is based on real-time data on human mobility/activity that provides an indirect proxy for infection transmission. One of the strengths of this approach is that, differently from epidemiological investigations, the data can be retrieved in real time and longitudinally, thus allowing to quickly capture possible changes in the most relevant settings for transmission. Furthermore, our approach could help minimize the noisy and biased data collection related to massive transmission events^28^. Yet, the approach used here is far from capturing all the finest details of human social contacts and thus the estimates on the contribution of different settings to SARS-CoV-2 transmission entail an unavoidable uncertainty.

To properly interpret our results, it is important to acknowledge the limitations of the assumptions included in our modeling exercise. First, we have considered a decrease of the transmission probability in outdoors as compared to indoors settings of 1/20^29^. Although this choice is guided by empirical evidence and our results are robust to this choice (see Supp. Material Section 7), further studies better quantifying the relative risk of indoor vs. outdoor transmission are warranted. Second, our model neglects to consider differences in the behavior that people follow when in contact with each other. It is indeed possible that contacts between relatives and friends have a larger chance of resulting in a transmission event as compared with interactions with strangers^30^. Third, we do not model nursing homes, which were severely hit by the COVID-19 pandemic across the globe. However, although they represent a key setting to determine COVID-19 burden in terms of deaths and patients admitted to hospitals and ICUs, they are possibly not central to capture the transmission dynamics of SARS-CoV-2 at the population level, which is the aim of this study. Although there is some location information from hospitals, we do not model them. Nonetheless, contact tracing studies from several countries have revealed that transmission within hospitals is relatively low, and hospital staff are more at risk from interactions with their coworkers (e.g. in the breakroom) or out in their communities^31, 32^.

In conclusion, the majority of NPIs introduced in large urban areas in March were effective to dramatically slow down the first wave of COVID-19 by greatly reducing the number of effective contacts in the population. Closing down schools, businesses, workplaces and social venues, however, took (and still does) an enormous toll on our economy and society. Our results and methodology allow for a real-time data-driven analysis that connects NPIs, human behavior and the transmission dynamic of SARS-CoV-2 to provide quantitative information that can aid in defining more targeted and less disruptive interventions not only at a local level, but also to assess whether local restrictions could trigger undesired effects at nearby locations not subject to the same limitations. Although nowadays the epidemiological landscape has dramatically changed by the introduction of vaccines, spread of more transmissible variants, and the build up of natural immunity, the results offered in this paper provide unique insights on the transmission pathways of SARS-CoV-2 and can be instrumental for the definition of location-based mitigation policies and for taking informed decisions about high-risk activities.

## Methods

We used individual-level mobility data of over half a million individuals distributed in New York and Seattle metropolitan areas during the months of February 2020 to June 2020 to estimate the day and type of venues where people might have interactions that yield to transmission events. To do that we extracted from the mobility data the stays (stops) of people in a large collection of around 440k settings^33^. With this information we built two synthetic populations, one for each metropolitan area, in which agents can interact in different settings: workplaces, households, schools, and the community (points of interest). We then explore the transmission of SARS-CoV-2 using a compartmental and stochastic epidemic model applied on top of this population.

The behavioral changes induced in the population by the introduction of several NPIs are naturally encoded in the mobility data, allowing us to characterize the effect of these interventions. We ran counterfactual simulations of our stochastic epidemic model to understand that effect. Furthermore, the resolution of this data allows us to characterize the spreading through different types of venues at different stages of the epidemic, depicting a complex picture in which the combination of both the characteristics of the place/setting and of the behavior of individuals who visit it determine its risk.

Lastly, the information about the statistical heterogeneity of the contact pattern of different individuals allows us to study the frequency and characteristics of behavior-related super-spreading events (SSE). We study the likelihood of finding a SSE per setting as a a function of time by looking at the number of infections produced by each individual in each location. A full description of the materials and methods is provided in the different sections of the Supp. Materials.

## Data Availability

The data that support the findings of this study are available from Cuebiq through their Data for Good program, but restrictions apply to the availability of these data, which were used under license for the current study, and so are not publicly available. Aggregated data used in the models are however available from the authors upon reasonable request and permission of Cuebiq. Other data used comes from the American Community Survey (5-year) from the Census, which is publicly available.

## Acknowledgements

Y.M. thanks M. Clarin for help with the design of Figure 1. N.E.D., I.M.L., MEH, and A.V. acknowledge the support of NIH/NIAID R56-AI148284. M.A., M.L., M.C. A.PyP. and A.V. acknowledge support from COVID Supplement CDC-HHS-6U01IP001137-01. M.C. and A.V. acknowledge support from Google Cloud Healthcare and Life Sciences Solutions via the GCP research credits program. A.V. acknowledge the support of McGovern Foundation and the Chleck Foundation. E.M. acknowledges partial support by MINECO (FIS2016-78904-C3-3-P and PID2019-106811GB-C32). Y.M. acknowledges partial support from the Government of Aragon and FEDER funds, Spain through grant E36-20R (FENOL), and by MINECO and FEDER funds (FIS2017-87519-P). A.A. and Y.M. acknowledge support from Banco Santander (Santander-UZ 2020/0274) and Intesa Sanpaolo Innovation Center. The funders had no role in study design, data collection, and analysis, decision to publish, or preparation of the manuscript.

## Author contributions statement

A.A., D.M-C., M.A., A.V., Y.M., and E.M. designed research; A.A. performed research with contributions from D.M-C. and M.B.; A.A., D.M-C., M.A., A.V., Y.M. and E.M analyzed the results. A.A. and E.M wrote the first draft of the manuscript; A.A., D.M-C., M.B., A.PyP., M.A., M.L., M.C., N.E.D., M.E.H., I.M.L., A.P., A.V., Y.M. and E.M. discussed results and edited the manuscript. All authors approved the final version.

## Additional information

### Competing interests

M.E.H. reports grants from the National Institute of General Medical Sciences during the conduct of the study; M.A. received research funding from Seqirus; A.V., M.C. and A.PyP report grants from Metabiota, Inc., outside of the submitted work. The authors declare no other relationships or activities that could appear to have influenced the submitted work.

## Code availability

The code is largely based on the one presented here https://github.com/aaleta/NHB_COVID.

## Notes

### Funding Statement

N.E.D., I.M.L., MEH, A.PyP. and A.V. acknowledge the support of NIH/NIAID R56-AI148284. M.C. and A.V. acknowledge support from Google Cloud Healthcare and Life Sciences Solutions via the GCP research credits program. E.M. acknowledges partial support by MINECO (FIS2016-78904-C3-3-P and PID2019-106811GB-C32). Y.M. acknowledges partial support from the Government of Aragon and FEDER funds, Spain through grant E36-20R (FENOL), and by MINECO and FEDER funds (FIS2017-87519-P). A.A. and Y.M. acknowledge support from Banco Santander (Santander-UZ 2020/0274) and Intesa Sanpaolo Innovation Center. The funders had no role in study design, data collection, and analysis, decision to publish, or preparation of the manuscript.

### Author Declarations

MIT Committee on the Use of Humans as Experimental Subjects. COUHES Exemption number #1812635935

### Summary of Updates

Updated version of the manuscript with new sensitivity and robustness tests of the methodology. Includes also minor revisions of the main text and supplementary material

