## Supplementary Material for "Quantifying the importance and location of SARS-CoV-2 transmission events in large metropolitan areas"

### Supplementary Material: Quantifying the importance and location of SARS-CoV-2 transmission events in large urban areas

**3 SARS-CoV-2 transmission model** **8**
**4 Calibration** **9**
**5 Effective reproduction number** **10**
**6 Superspreading events** **10**
**7 Sensitivity analysis** **10**
**References** **24**

### 1 Mobility data

The mobility data was obtained from Cuebiq, a location intelligence and measurement company. The dataset consists of anonymized records of GPS locations from users that opted-in to share the data anonymously in the New York metropolitan area over a period of 5 months, from February 2020 to June 2020. In addition to anonymizing the data, the data provider obfuscates home locations to the census block group level to preserve privacy. Data was shared in 2020 under a strict contract with Cuebiq through their Data for Good program where they provide access to de-identified and privacy-enhanced mobility data for academic research and humanitarian initiatives only. All researchers were contractually obligated to not share data further or to attempt to de-identify data. Mobility data is derived from users who opted in to share their data anonymously through a General Data Protection Regulation (GDPR) and California Consumer Privacy Act (CCPA) compliant framework.

Our sample dataset achieves broad geographic representation for our two populations, in the New York and Seattle metropolitan areas, defined as the Core Based Statistical Areas (CBSA) by the US Census[1]. CBSA are areas that are socioeconomically related to an urban center. This provides a self-contained metropolitan area in which people move for work, leisure or other activities. Some of the CBSAs we consider span several states. For example the New York CBSA contains areas of the state of Connecticut, New Jersey, Philadelphia, and New York. We filter all anonymous devices which were not observed each month, in order to make sure we had a stable population with enough granularity and representativeness of agents over the whole period. The population and number of anonymous devices detected in the real data by census area are highly correlated for both census county subdivision regions, with a $\rho = 0.796$ (Pearson correlation) with a CI between 0.783 and 0.807 for the New York region, and a $\rho = 0.948$ (Pearson correlation) with a CI between 0.937 and 0.957 for the Seattle region. We built such correlations between the population for each county subdivision and the number of devices in our dataset. Despite these large correlations our mobility dataset has a small income bias towards areas of higher income, specially in the NY metro area. However, as shown in Supp. Section 7, our results do not depend on that bias.

#### 1.1 Points of Interest

Fousquare Public API was used to retrieve a large collection of (Points of Interest) POIs in the NY and Seattle metro areas. Although Foursquare data is also crowd-sourced resource, it exhibits some editorial control. Their database of POI not only comes from users of their Swarm (check-in) platform, but is built by aggregating data over 46k different trusted sources[2]. Several studies confirm that although none of the POI databases is complete, Foursquare is one of the best in number of POIs, location accuracy and number of categories represented[3].

We use a dataset of 375k Points of Interest (POI) in the New York metropolitan area and 70k Points of Interest in Seattle metropolitan area collected using the public Foursquare API. Those POIs are categorized using the Foursquare taxonomy of places which has ten main categories. There are also 638 subcategories, see[4] for a complete list of them. We manually curated every subcategory in the taxonomy to be reassigned to twelve new principal categories: Arts & Museums, College, Entertainment, Exercise, Food & Beverages, Grocery, Health, Other, Outdoors, School Service, Shopping and Transportation. In our database the New York metropolitan they are distributed as follow Art & Museum (2.1%), College (2.9%), Entertainment (7.6%), Exercise (2.8%), Food & Beverage (17.7%), Grocery (2.6%), Health (7.5%), Other Places (13.1%), Outdoors (8.2%), School (2.3%), Service (16.6%), Shopping (8.3%), Sport & Events (0.6%) and Transportation (6.9%). For the Seattle metropolitan area POIs we have 69,906 POIs that are distributed as follows Art & Museum (2.7%), College (2.3%), Entertainment (7.1%), Exercise (2.7%), Food & Beverage (14.5%), Grocery (2.1%), Health (8.1%), Other Places (15.1%), Outdoors (7.8%), School (1.6%), Service (18.2%), Shopping (8.3%), Sport & Events (0.8%) and Transportation (7.8%). Despite our dataset contains many venues and places which are companies or business, some evidence that our dataset covers most of the public places comes by comparing them to official statistics: for example, we have 2,155 art galleries in the NY metro area compared to the 1,500 estimation for NY City only. On the other hand we have 9,810 groceries in the NY metro area in our POI database which compares quite well with the 11,791 grocery business reported by the U.S. Bureau of Labor Statistics in their Quarterly Census of Employment and Wages in the NY Metro area[5].

#### 1.2 Stays

From the combination of the mobility data and the POIs we extract "stays", as the unique places where anonymous users stayed (stopped) for at least 5 minutes. Each device frequently broadcast its location to a central server by sending its latitude, longitude, device ID, and the exact date and time of the event. When a person spends significant time at a single location, measurement uncertainty will cause a number of events to be scattered around the actual location. To map these events to a single stay with an accurate time and location, we use the Infostop algorithm[6]. First, to extract the locations of stays, the algorithm clusters consecutive events together if the locations are less than 25 meters apart. The location of this cluster is computed by taking the median of the latitudes and longitudes. Moreover, to better estimate the location of places that are visited frequently by the same user, the algorithm also checks whether different clusters appear within 25 meters of each other and assigns a single consistent location to all connected clusters by recomputing the median latitude and longitude. Finally, a stay is registered whenever at least two subsequent events are registered at one of these locations where the first and last event respectively mark the start and end time of the stay. The minimum duration of a stay is set to 5 minutes to make sure we are only including actual contact between people instead of people that, for example, pass each other on an intersection.

Some of the stays happen within or close to places (Points of Interest). We attributed a stay to the closest POI up to a distance of 50m, otherwise that stay is discarded. We do not make this attribution if the closest place is further than 50meters (see also the SI for a sensitivity analysis with other maximum distance to POIs). Although we use 50m as an upper bound, in reality the average distance to the attributed POI is smaller, 19.43 meters on average in the metro areas of NY and Seattle, which is smaller than the average distance between nearest POIs, 33 and 39 meters respectively. In areas with large numbers of POIs like Manhattan, the distance to the attributed closest POI is even smaller. Note that we attribute each stay to a single POI and in turn, to a single category of place. We have also checked that our results do not depend significantly on the 50 meters threshold for the attribution of the stays (see Supp. Section 7). Stays are then aggregated at place level.

For privacy reasons, our data is obfuscated around home and workplaces to the level of Census Block Groups. Thus the attribution between the mobility data and home and workplaces happen at the level of Census Block groups and not specific POIs. We estimate the home Census Block Group of the anonymous users as the one in which they are more likely located during nighttime. This results in a dataset of the places people stayed including the POIs in the community layer, the CBG of their workplaces that anonymous users visited, and the most likely census block group of where the device owner lives.

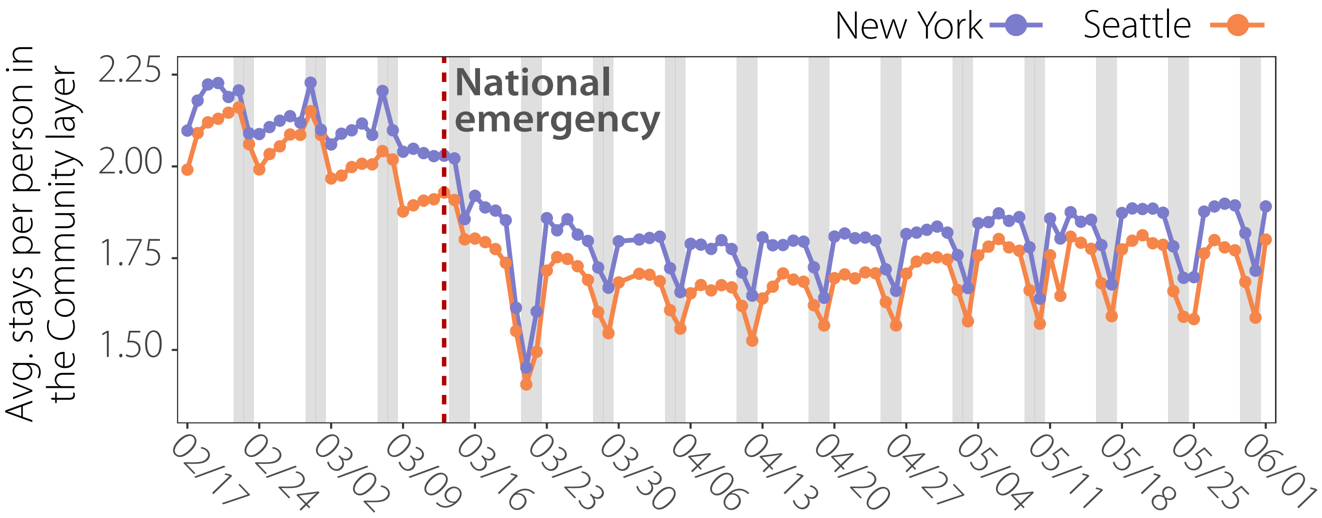

**Figure S1.** Evolution of the average number of stays in the Community layer per observed person for New York and Seattle metropolitan areas. Vertical red dashed line indicates when National Emergency (N.E.) is established.

In Figure S1 we can see the daily evolution of the average number of stays per observed person for New York and Seattle only in the community layer. Also in Figure S2 we can see the total observed number of stays in our datasets. Two weeks before we can see that Seattle started to see a small change in the mobility behaviour, however, for New York City we can start to see that pattern one week before the national emergency. The average number of daily stays per agent for New York before the N.E. is 2.14 with a 95% CI [2.12, 2.17]. On the other hand, for Seattle is 2.05 with a 95% CI [2.02, 2.08]. After the national emergency there is an abrupt decrease for both cities in the number of stays (see Figure S2). Two weeks after the national emergency the average number of stays per person stabilized and starts to an slightly and steady increase. Eleven weeks after the national emergency, the average number of stays per person has recovered slightly, but it did not recover its basal state for both cities. The average number of daily stays per observed agent for New York after the N.E. is 1.83 with a 95% CI [1.81, 1.84]. On the other hand, for Seattle is 1.72 with a 95% CI [1.71, 1.74].

We can see in Figure S2 the daily evolution of the total number of stays to each category and their fraction distribution. Figure S2 (a) for New York and (c) for Seattle represent the total number of stays at the community layer, we can see a similar pattern as in in Figure S1 (a) before and after the national emergency. Figure S2 (b) for New York and (d) for Seattle show normalized number of stays. We can see a reduction of non-essential places after the national emergency due to the social distancing policies.

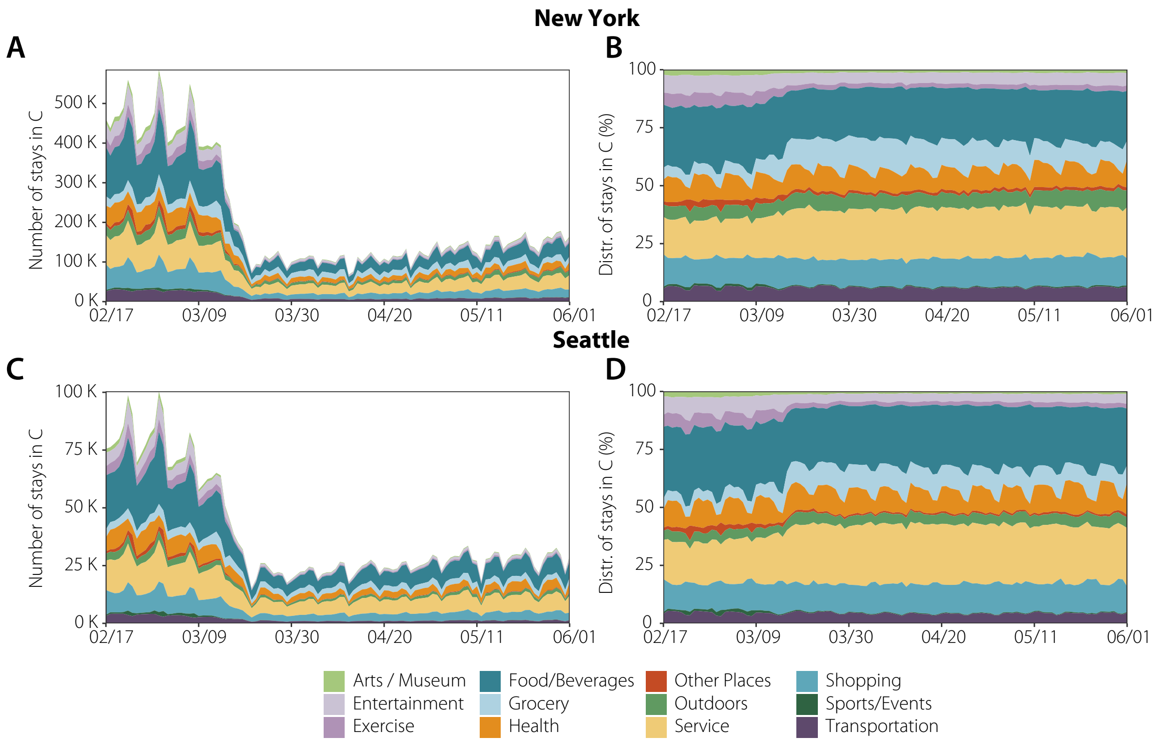

**Figure S2.** The comparative evolution of the number of stays (left) and distribution (right) of stays in the Community layer for the different metropolitan areas, New York (top) and Seattle (right).

Finally, in Figure S3, we can see the comparison of the average time per stay for each city and category before and after the national emergency. There is a significant decrease in time spent per stay for nearly each category in both cities. However, the grocery and the transportation categories are those with the smallest change in the average time for both cities. Moreover, the shopping category does not barely change in New York, but it does in Seattle. On the other hand the Food & Beverages category decrease in New York, but it does not in Seattle.

### 2 Network structure

#### 2.1 Agents

Our population consists of two different sub-populations, adults and children. Adults are sampled from anonymous individuals in the mobility data collected by Cuebiq, each adult is associated with a home location assigned to a US Census block group which is provided by our location data provider. We used those anonymous individuals to construct synthetic populations by assigning them different socio-demographics using highly detail macro (census) and micro (survey) data. This procedure to create synthetic representative households and demographic traits is documented in[7].

Following this process we generate two synthetic populations, one for the New York metropolitan area and the other one for the Seattle metropolitan area. The New York synthetic population consists of 565k agents (3.0% of the population in the New York metropolitan area), 78.02% of them are adults and 21.98% are children. Distribution of age groups are shown in Figure S4a where we can see the that our synthetic population age distribution compares well against the US census data. The same happens for the household size distribution, where 31% of the households are of size two, 29.5% of size one and the rest are of size three or bigger, see Figure S4b. The Seattle synthetic population consists of 106k agents (2.9% of the population in the Seattle metropolitan area) with 76.7% of them adults and 23.3% are children. The age groups distribution is shown in Figure S4c where we can see that they compare well with the demographic distribution. Household size distribution is very similar to the NY metro area, with 27.2% of size one, 34.8% of size one and the rest of size three or bigger. In Figure S4d we can see the comparison of our synthetic households population distribution against the US census data.

The population that we are using to build the contact matrices is statistically representative of the total population in the urban areas. Previous work has shown that this sample of the population can accurately describe the number of visits, lifestyles and the mobility of the whole population[8], once it is re-scaled using post-stratification methods. Given that we re-scale the

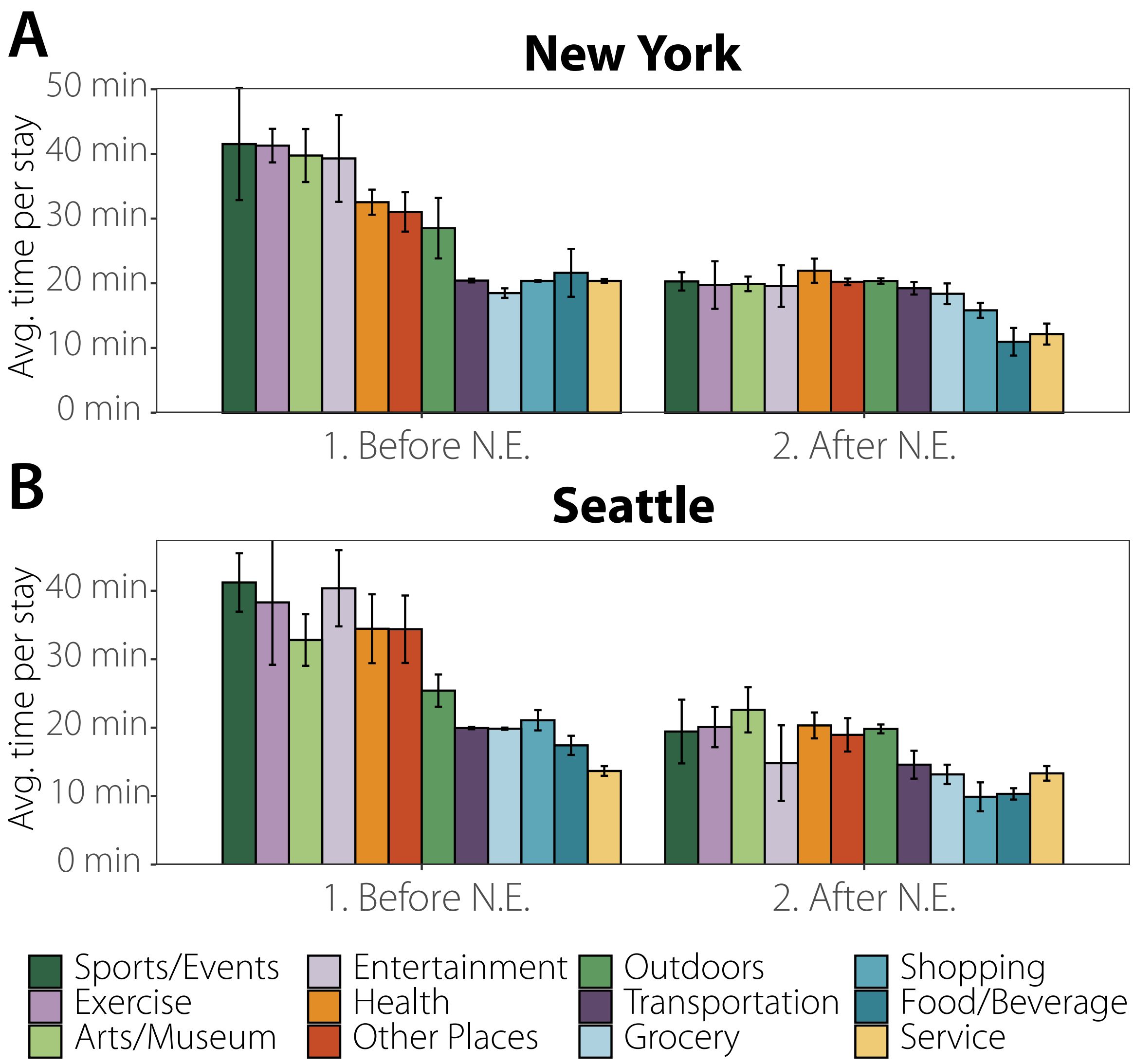

**Figure S3.** Average time per stay for each place category before and after the National Emergency (N.E.) for (a) the New York Metropolitan Area and for (b) the Seattle Metropolitan Area.

transmissibility and the number of effective contacts to reproduce effectively the dynamics of the infection at the population level, we believe that our results do not depend on the size of our sample

##### 2.2 Contacts

Visits to different POIs were used to estimate probabilistically the contacts between anonymous users. Not that our estimation of contact between individuals is not a direct observation of colocation events. Although the mobility dataset we use is large, colocation events between individuals are still quite sparse. Because of this sparsity, and to protect individual privacy in our analysis, we have adopted a probabilistic approach to measure co-presence (and probability of transmission) in all locations mapped in the dataset. Our objective is to build the contact matrix $\omega_{ij}$ between individuals $i$ and $j$ using those estimations of co-presence in the different layers where those contacts are possible, Home, Schools, Workplace, and Community.

In order to explain better our approach let us consider the homogeneous mixing approach in a contact network perspective. We assume to have $N$ individuals who are homogeneously mixed. This implies that each individual is potentially in contact with anybody else. Thus, we have a connection $\omega_{ij} = 1$, among each pair of nodes. Then, the rate of contacts $c_i$ for the individual $i$ is $c_i = \sum_j m\,\omega_{ij} = m(N-1)$, where $m$ is an appropriate factor ensuring that the number of average effective contacts per

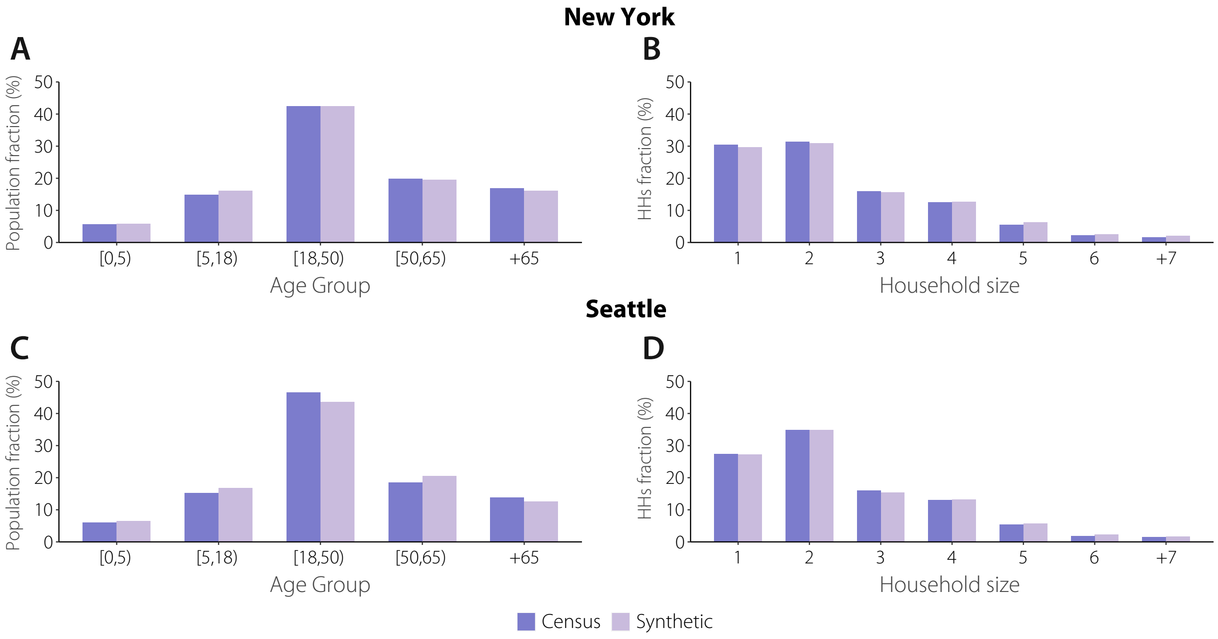

**Figure S4.** Age groups and households demographics compared against the US Census data. (a) Age groups distribution and (b) households size distribution for the New York Metropolitan Area. (c) Age groups distribution and (d) households size distribution for the Seattle Metropolitan Area.

individual unit time in the system is equal to $\kappa$. Hence,

$$\kappa = N^{-1}\sum_i c_i = N^{-1}\sum_{i,j} m\,\omega_{i,j} \tag{1}$$

yielding

$$m = \frac{\kappa}{N^{-1}\sum_{ij}\omega_{ij}} = \frac{\kappa}{N-1} \tag{2}$$

This provides the usual expression for the rate of contact $\omega'_{ij} = \kappa/(N-1)$, that is multiplied by the transmissibility per contact $\alpha$ to give the rate (or probability) of infection per contact. This finally leads to the force of infection of a susceptible as

$$P_{S\to I} = 1-(1-\frac{\alpha\kappa}{N-1})^I = 1-(1-\frac{\beta}{N-1})^I \simeq \frac{\beta I}{N},$$

where $\beta = \alpha\kappa$ is the transmissibility used in homogeneous model and the last approximations is valid for very large $N$.

In order to go beyond the homogeneous assumption, from our data we can consider that individuals who are never visiting the same places are never in contact. This is additional information of which we are certain. So for each individual we can list each of the places p that they visit and assume that we can have a link between two individuals if they have the same place in their list $\omega^p_{ij} = \delta_{i,p}\delta_{j,p}$, where $\delta_{i,p} = 1$ if the place $p$ is on the list of visited places of individual $i$ and zero otherwise. This step improves on the homogeneous assumption as it rules out possible contacts among individuals that can never meet. Further we can consider that the potential contacts among individuals is larger for individuals that can meet in more than one place. We can then define $\omega_{i,j} = \sum_p \omega^p_{i,j}$ , thus considering that some individuals have more potential contacts. It is worth remarking that we are still considering that each potential contact has the same weight as in the homogeneous assumption. In order to define properly the contact rate/probability per unit time we need to use Eq. (1) thus defining

$$m = \frac{\kappa}{N^{-1}\sum_{i,j}\omega_{ij}} = \frac{\kappa}{\langle\omega_{ij}\rangle} \tag{3}$$

where we defined $\langle\omega_{i,j}\rangle$ as the average weighted contacts among individuals. This yields the effective rate of contact among individuals $i$ and $j$ as

$$\omega'_{ij} = \frac{\kappa\sum_p \delta_{i,p}\delta_{j,p}}{\langle\omega_{ij}\rangle} \tag{4}$$

In order to improve further on this approach we can consider that places are not visited in a deterministic way. This implies that each individual has a probability to visit a specific place that is $1/n_{i,p}$, where $n_{i,p}$ is the number of places visited by the individual $i$ in a given period. We can therefore define

$$\omega_{ij} = \sum_p \frac{1}{n_{i,p}} \frac{1}{n_{j,p}}. \tag{5}$$

This approach still considers potential contacts only among individuals however with a weight that depends on the variability of places of each individual. As before the rate/probability of contact would be:

$$\omega'_{ij} = \frac{\kappa \sum_p n_{i,p}^{-1} n_{j,p}^{-1}}{\langle \omega_{ij} \rangle} \tag{6}$$

So far we did not consider at all the time spent in each location. We can therefore improve on the probability to be in a place by weighting the number of places $n_{i,p}$ by the time spent on average in each place. This finally leads to the expression:

$$\omega_{ij} = \sum_p \frac{T_{i,p}}{T_i} \frac{T_{j,p}}{T_j} \tag{7}$$

where $T_{i,p}$ is the time spent by individual $i$ at location $p$ and $T_i$ is equal to the sum of all time spent in places in the community by individual $i$. In this case the rate of interaction will be:

$$\omega'_{ij} = \frac{\kappa \sum_p \frac{T_{i,p}}{T_i} \frac{T_{j,p}}{T_j}}{\langle \omega_{ij} \rangle}. \tag{8}$$

This is the expression we use in our work. It is important to stress that this expression is improving on the homogeneous assumption as it considers that effective contacts can occur only in places visited by both individuals, and considers that each contact is weighted by the probability for each individual to be in that place. The approach however does not account for concurrency of visits. In this respect it is still adopting an homogeneous perspective in that all places visited at any time corresponds in a potential contact.

The next steps to improve on this approach would be indeed to consider concurrency of visits. It is thus tempting to consider that each contact is weighted by $T_{i,p}/T$, where T would be the specific amount of time of the day. One could assume the 8 hours of the working time or the 24 hours cycle of the day. This is a tempting solution but introduces a number of issues. For instance the time that should be considered in the normalization depends on the places. For instance restaurants have specific bracket of times during the day, and concurrency should be evaluated on specific hours of the day and specific days (for instance the week- end). The same is for places like movie theatres, museums etc. Furthermore, during the lockdown the concurrency normalization should all be re-evaluated to be consistent in their definition as the number of hours in the community of the population has drastically changed. In other words, we are not sure if the simple normalization by a fixed number of hours although trying to capture the concurrency of contacts is actually introducing unwanted and uncontrolled biases. For this reason we decided to work with the approach of Eq. (8), for which all the assumptions can be clearly stated and provides an obvious improvement with respect to the fully homogeneous assumption.

Using our probabilistic approach to detect contacts, we build our contact network in each of the layers::

- **Community weighted contact network**. In the community layer contacts are built by estimating probabilistically the interaction between two individuals who visit the same POI. Specifically, the weight, $\omega_{ijt}^C$, of a link between individuals $i$ and $j$ within the community layer at day $t$ is computed according to the expression:

$$\omega_{ijt}^C = \sum_p^n \frac{T_{ipt}}{T_{it}} \frac{T_{jpt}}{T_{jt}}, \qquad \forall i,j \tag{9}$$

  where $T_{ipt}$ is the total time that individual $i$ was observed at place $p$ in day $t$ and $T_{it}$ is the total time that individual $i$ has been observed at any place set within the community layer that day $t$. The distribution of values of $\omega_{ijt}$ is very broad. For example in NY $\omega_{ijt}$ as a mean of 0.395, a median of 0.279 and 25% and 75% quantiles of 0.095 and 0.65238 respectivelly. Finally, for robustness and computational reasons, we have included only links for which $\omega_{ijt}^C > 0.01$, removing 2.88% of the original links. For other values of the threshold like $\omega_{ijt}^C > 0.005$ and $\omega_{ijt}^C > 0.02$ we would remove 1.19% and 6.19% of the links respectively. Note however that since those links have very small weights, our results for the epidemic spreading do not depend significantly of the threshold chosen provided that it is small.

- **Workplace weighted contact network**. For privacy reasons, our data is obfuscated around home and workplaces to the level of Census Block Groups. To get a proxy of contacts at the workplace, we assume that all workers in the same Census Block Groups have a probability to interact. To account for the potential number of working places in that area, we weight that probability by the number of POIs at the same census block group. Therefore, the contact weight, $\omega^W_{ijt}$, of a link between individuals $i$ and $j$ within the same workplace at day $t$ is given by:

$$\omega^W_{ijt} = \sum_{\alpha \in \text{CBG}} \sum_{\beta \in POI(\alpha)} \frac{\delta_{i\alpha t}}{N_{POI}(\alpha)} \frac{\delta_{j\alpha t}}{N_{POI}(\alpha)} = \sum_{\alpha \in \text{CBG}} \frac{\delta_{i\alpha t}\delta_{j\alpha t}}{N_{POI}(\alpha)}, \qquad \forall i,j \tag{10}$$

where $POI(\alpha)$ is the set of POIs in the census block group $\alpha$, $N_{POI}(\alpha)$ is the number of POIs in $\alpha$, $\delta_{i\alpha t}$ is the binary variable of observing or not an individual at her workplace within census block group $\alpha$ at day $t$. As before, we have included only links for which $\omega^W_{ijt} > 0.01$.

**2) Household weighted contact network**. We first identify individuals' approximate home place as their most likely visited census block group at night. Then we assign a synthetic representative household and demographic traits as documented in[7]. To assign weights, we assume that the probability of interaction within a household is proportional to the number of people living in the same household (well-mixing). Therefore, the weight, $\omega^H_{ij}$, of a link between individuals $i$ and $j$ within the same household is given by:

$$\omega^H_{ij} = \frac{1}{(n_h - 1)} \tag{11}$$

where $n_h$ is the number of household members. This fraction is assumed to be the same for all individuals in the population. We assume this layer is static throughout our period.

- **School weighted contact network**. To calculate the weights of the links at the school layer, we mix together all children that live in the same census tract. Interactions are considered well-mixed, hence, the probability of interaction at a school is proportional to the number of children at the same school. Therefore, the weight, $\omega^S_{ij}$, of a link between children $i$ and $j$ within the same school is given by:

$$\omega^S_{ij} = \frac{1}{(n_s - 1)} \tag{12}$$

where $n_s$ is the number of school members. This layer is removed on March 16 in both metropolitan areas to account for the imposed school closure.

To calibrate the relative importance of each layer in the spreading process we further multiply the weights by their corresponding $\kappa$. In particular, with $\kappa = 4.11$ in the household layer, $\kappa = 11.41$ in the education layer, $\kappa = 8.07$ in the workplace layer and $\kappa = 2.79$ in the community layer[7], see Eq. (8)

#### 3 SARS-CoV-2 transmission model

To model the natural history of the SARS-CoV-2 infection, we implemented a stochastic, discrete-time compartmental model on top of the contact network $\omega_{ijt}$ in which individuals transition from one state to the other according to the distributions of key time-to-event intervals (e.g., incubation period, serial interval, etc.) as per available data on SARS-CoV-2 transmission. In the infection transmission model, susceptible individuals (S) become infected through contact with any of the infectious categories (infectious symptomatic (IS), infectious asymptomatic (IA) and pre-symptomatic (PS)), transitioning to the latent compartment (L), where they are infected but not infectious yet. Contacts between infected and susceptible individuals depend on the contact network estimated for each day. Thus, the probability that a susceptible node $i$ is infected by an infectious node $j$ in infectious compartment $type$ and location $l$ is:

$$P(S_i + I_j \to L_i + I_j) = 1 - e^{-\beta_{type} w_{i,j,l}(t)\Delta t} \tag{13}$$

where $\Delta t = 1$ day. Latent individuals branch out in two paths according to whether the infection will be symptomatic or not. We also consider that symptomatic individuals experience a pre-symptomatic phase and that once they develop symptoms, they can experience diverse degrees of illness severity, leading to recovery (R) or death (D). The value of the basic reproduction number is calibrated to the weekly number of deaths.

The values of all the disease parameters used for simulating the transmission dynamics are given in Table S1. Figure S5 shows the numerical distributions of these parameters as resulting from simulations of the model, computed for the case of New York with $R_0 = 3.2$ (see Supp. Section 4).

| Parameters | Description | Age group | Value | Ref. |
|---|---|---|---|---|
| $r$ | relative infectiousness of asymptomatic individuals | - | 50% | † |
| $k$ | proportion of pre-symptomatic transmission | - | 50% | [9] |
| $\varepsilon^{-1}$ | incubation period (gamma distributed) | - | shape = 2.08<br>rate = 0.33 | [10] |
| $p$ | proportion of asymptomatic | - | 40% | [9] |
| $\gamma^{-1}$ | pre-symptomatic period | - | 2 days | [11] |
| $\mu^{-1}$ | time to isolation | - | 2.5 days | |
| $\delta^{-1}$ | days from isolation to death | - | 12.5 | [9] |
| IFR | infection fatality ratio | 0-9<br>10-19<br>20-29<br>30-39<br>40-49<br>50-59<br>60-69<br>70-79<br>$\geq 80$ | 0.00161%<br>0.00695%<br>0.0309%<br>0.0844%<br>0.161%<br>0.595%<br>1.93%<br>4.28%<br>7.80% | [12]‡ |
| $T_n$ | Notification of death | - | 7 days | [9] |
| $\theta$ | outdoor transmissibility | - | 0.05 | [13] |

**Table S1.** Baseline set of parameters. †: assumed ;∗: calibrated to the generation time $T_g$; ‡ Only applied to symptomatic individuals. As such, a correction factor of 1/(1-p) is applied to all age groups.

#### 4 Calibration

The model has two free parameters: (1) the number of infected individuals in each city on the first day for which we have data to build the interaction networks (02/17/2020) and (2) the value of $R_0$.

The first date contained in our data is 02/17/2020, a time when it is estimated that there were already several infected individuals in New York. In particular, we use the estimates provided by the GLEAM model[14]: 292 latent individuals in New York City and 39 in Seattle. To initialize the system into such a state one could select that number of agents randomly from the simulation and move them into the latent compartment. However, this would not resemble the real evolution of the epidemic, which does not infect people at random but instead follows the path imposed by the behavior of individuals. For this reason, we initialize the system with 1 infected individual and run the model in a loop using the networks of the first week available (02/17/2020 to 02/23/2020). Once the estimated number of individuals is observed, the system starts to run in calendar time from 02/17/2020 to 06/01/2020 (each step corresponds to 1 day). This allows us to start the simulation with the estimated number of latent individuals in each city on 02/17/2020 without having to select them at random.

We use an Approximate Bayesian Computation (ABC) rejection algorithm to obtain the posterior distribution of $R_0$. We sample the transmissibility from a uniform prior so that $R_0$ is in the range 1.5 to 4.5 and compare the output of the model with the weekly estimated number of deaths as a consequence of COVID-19 for each city[15]. The obtained posterior distribution $P(R_0 = x|E)$ is shown in Figure S6. The estimation of $R_0$ as a function of the transmissibility is performed using the expression proposed in[16]:

$$R_0 = \frac{r}{\sum_{i=1}^{n} y_i (e^{-ra_{i-1}} - e^{-ra_i})/(a_i - a_{i-1})} \tag{14}$$

where $r$ is the growth rate and $y_i$ and $a_i$ represent the frequency and the bins of the histogram representation of the generation time extracted from the simulation.

In Figure S7, we show the fitting of the model presented in the main text but without the curves corresponding to New York to enhance the readability of Seattle curves.

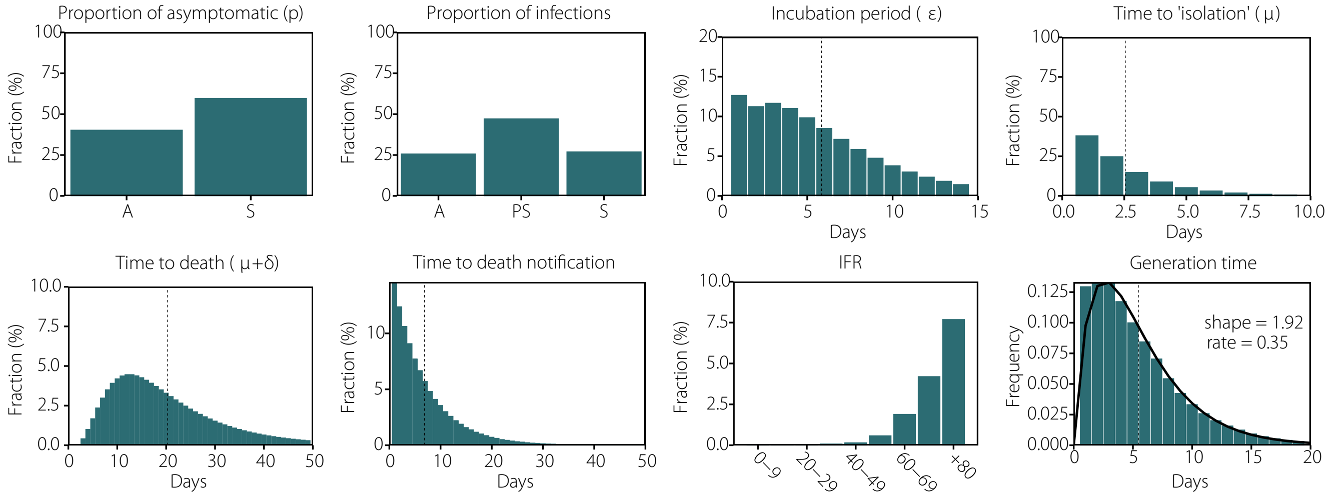

**Figure S5.** Numerical distributions of the model parameters extracted from the simulations performed for New York with $R_0 = 3.2$. The generation time distribution is well fitted by a gamma distribution with shape = 1.92 and rate = 0.35. The three infectious compartments, asymptomatic, symptomatic and pre-symptomatic individuals are labeled as $A$, $S$, and $PS$, respectively.

#### 5 Effective reproduction number

The effective reproduction number can be estimated using case count data as reported by the authorities. We relied on the technique proposed by Zhang et al.[17], but a review of other techniques can be found in[18] . In[19], the authors estimated this quantity for different areas of the world. As we show in Figure S8, $R(t)$ in our model drops below 1 on the same date as in the estimated $R(t)$ from real data, signaling that the peak occurs at the same time in both.

#### 6 Superspreading events

In heterogeneous populations it is possible for an infected individual to produce an usually large number of secondary cases. This is known as a super-spreading event (SSE). To define a SSE we follow Lloyd-Smith et al[20]:

1. Estimate the effective reproduction number, $R$
2. Compute a Poisson distribution with mean $R$
3. Define a SSE as any infected individual who infects more than the 99-th percentile of the Poisson distribution within a certain category of place.

In Figure S9 we test the hypothesis of the 20/80 rule according to which 20% of the infected individuals produce 80% of the infections. Note that this does not imply that said 20% of individuals are super-spreaders. In fact, the large majority of them do not produce any secondary infections, inline with what has been observed in highly detailed empirical studies[21].

In Table S2 we report the probability of having a SSE within each category before and after the declaration of the National Emergency. We observe a drastic reduction of the probability after 03/13.

#### 7 Sensitivity analysis

##### 7.1 Distance to POIs

While constructing the network, we attributed a stay to a given POI if it was no further than 50 meters from the POI center. In this section we test more strict conditions for that attribution, i.e. a threshold of just 10 meters. Note that this more strict condition for attribution lowers the number of potential visitors to the POI but also lowers the distance between people in the venue, making physical contact more likely. In Figure S10 we show the results for this scenario.

A more restrictive definition of stay yields a much sparser network in the community layer, while it does not affect the rest of the layers. We can see that to obtain the observed number of deaths under these conditions, the fraction of infections attributed to the workplace layer is increased. Nevertheless, the distribution of infections across settings is fairly similar, signaling that the results are robust to this definition.

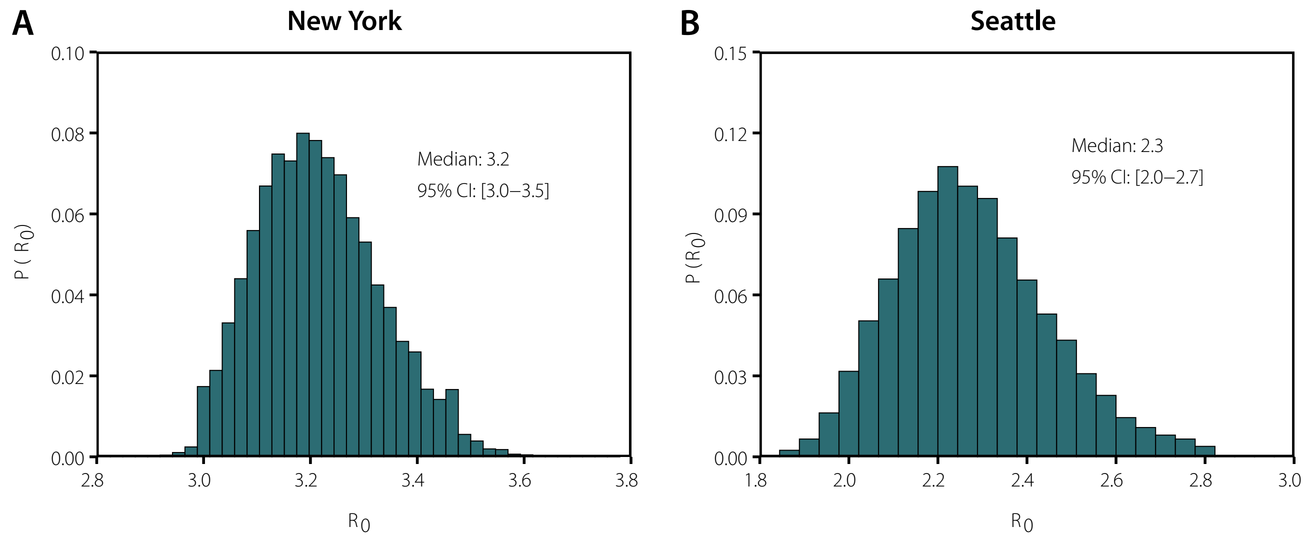

**Figure S6.** Posterior distribution of $R_0$ given the number of weekly deaths in each region as evidence.

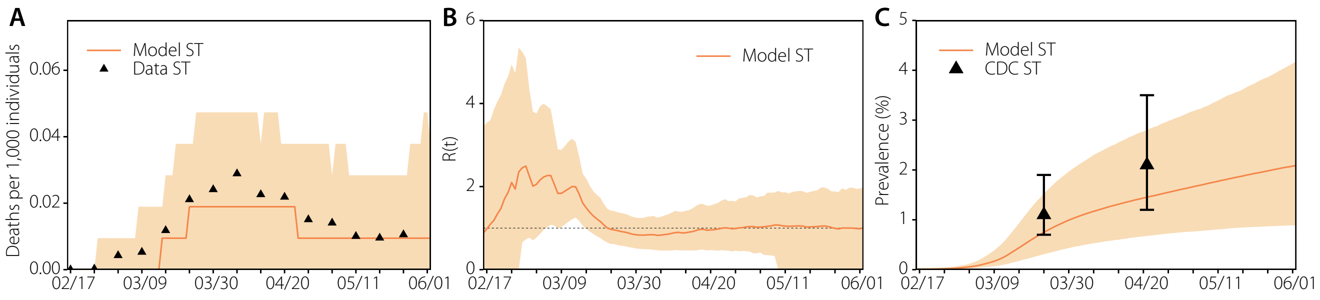

**Figure S7.** Model fit to Seattle data as reported in the main text.

##### 7.2 Model parameters

To test the dependency of the results with the values assumed in the model, we have explored three different scenarios: larger transmissibility during the pre-symptomatic phase ($k = 0.75$), Figure S11; longer time from death to notification ($T_n = 14$ days), Figure S12; and larger outdoor transmission ($\theta = 0.10$), Figure S13. The results are consistent in all cases with only slight variations on the value of $R_0$.

##### 7.3 Behavioral changes

The aggregated change in behavior due to the evolution of the epidemic as well as the introduction of non-pharmaceutical interventions is already contained in the mobility data. This leads to the sudden drop in the number of contacts following the declaration of the National Emergency. However, at the individual level, it might be possible that some individuals in the dataset lowered their contacts due to having developed symptoms, even if in our simulations they do not get infected at all and vice versa. But for anonymity reasons, it is not possible to relate the medical history of individuals and our agents and, thus, we cannot know the reason why an individual might have changed her behavior. From the point of view of the individual this observation is important, but since we are working on aggregated metrics this observation does not affect the results.

To demonstrate this, in Figure S14, we show the results in which we completely remove symptomatic transmission. This extreme scenario would represent a situation in which every time an individual develops symptoms, she gets completely isolated. As we can see, the overall results are close to the ones we have presented so far. The reason is that our model is fitted to the number of deaths and, thus, the total number of infections is fixed (as a function of IFR). If we remove one type of transmission, then the transmissibility of the other types has to be increased to achieve the same number of deaths, yielding similar results.

##### 7.4 Economic and age bias

The complete sample of users is slightly biased towards higher income individuals. Specifically, the penetration ratio (number of mobile phone users to adult population) in each census tract is correlated with the median household income, $\rho = 0.28 \pm 0.02$

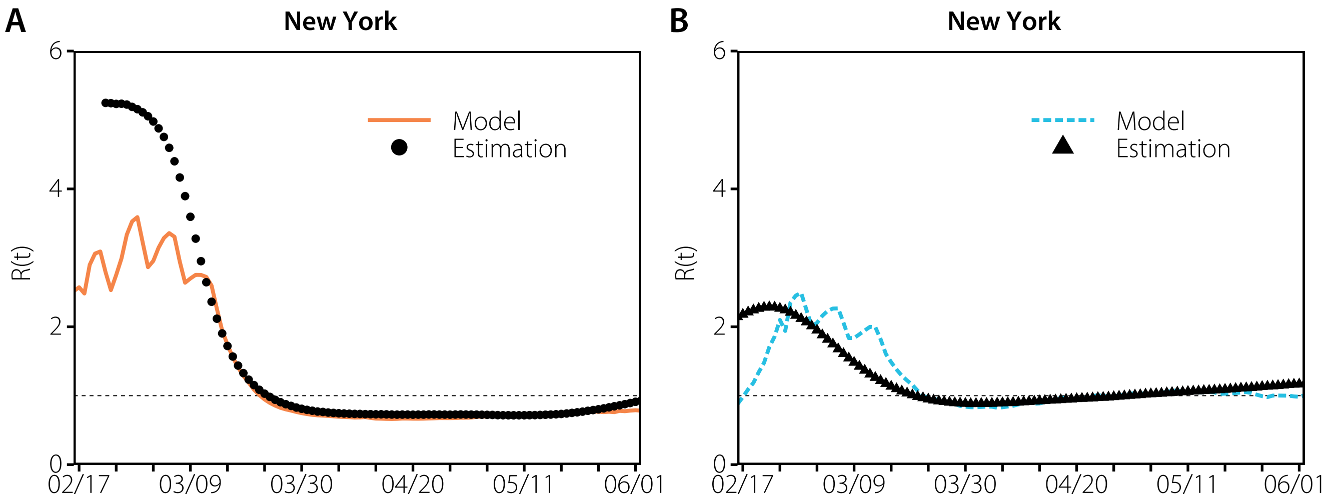

**Figure S8.** Effective reproduction number in both areas as obtained by our model and estimated by[19].

in NY and $\rho = 0.18 \pm 0.02$ in Seattle metro areas. However the correlation of the penetration ratio with the number of people above 64 years old in each census tracts is small $\rho = 0.17 \pm 0.04$ in the NY area or not significant $\rho = -0.06 \pm 0.11$ in the Seattle area. To analyze the impact of this bias, we have investigated the dynamics of our model in a different set of users obtained by downsampling each economic groups (median income quartiles in each metro area) to have a better representation of them. In Figure S15 we report the results obtained using this new sample of users. As we can see, the results remain largely unaltered, signaling that the distribution of contacts per type of venue is not affected by this bias.

##### 7.5 Removing deaths in long term care facilities

We have tested the sensitivity of the results if we remove deaths produced in long term care facilities from the total amount of deaths used to fit the model, Figure S16. We observe that the overall behavior is the same, although the number of total infections required is smaller, yielding a lower prevalence.

##### 7.6 Longer stays

We have tested the sensitivity of the results with a more strict definition of stay (minimum 15 minutes instead of 5 minutes), Figure S17. We observe a slight increase in the Arts & Museums category before the declaration of the National Emergency, and one in the Grocery category after the declaration. This indicates that individuals tended to stay for longer in groceries in this period, but the rest of the results remain largely unaffected.

##### 7.7 Differential age-susceptibility

There is preliminary evidence that children and adolescents have lower susceptibility to SARS-CoV-2[22]. In figure S18, we report the results when children and adolescents younger than 20 years have an odds ratio of 0.56 for being an infected contact compared with adults. The main difference with the previous scenarios is that the fraction of infections in the school layer is lower, but that does not have any impact on the rest of the results.

##### 7.8 Larger household transmissibility

We analyze the effect of increasing the within household transmissibility after the declaration of the N.E. In particular, we increase said transmissibility by 50% to account the extra time that individuals stay in the household. In figure S19, we show that this produces an increase in the infections within the household layer during this period, but the rest of the results remain largely unaltered.

##### 7.9 Different weight choice

To properly calibrate the relative importance of contacts in each layer it would be necessary to know the exact empirical contribution of each setting to the total number of infections. This is something completely unknown to this date, with estimates that vary widely across regions and time. Indeed, since currently the only way to empirically obtain the information is through surveillance systems, this task is very prone to errors, especially when cases are high, as in the scenario we are considering in this manuscript. Furthermore, any intervention will modify the relative contributions, limiting the applicability of the results.

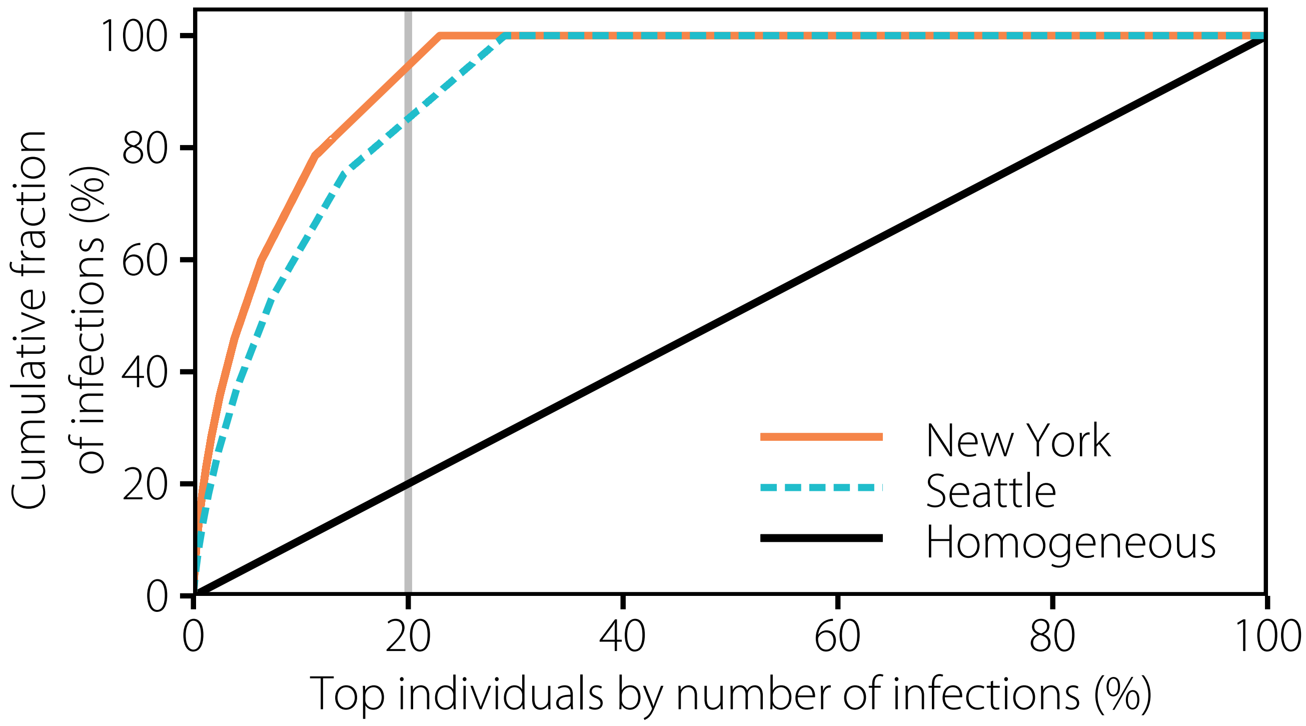

**Figure S9.** Individuals are ranked according to the number of infections they produce. The cumulative fraction of infections found in both cities is compared with the one that would be obtained in a completely homogeneous system.

For this reason, we have relied on the distribution of contacts across layers that has been estimated through multiple surveys in a wide range of countries as a proxy to the relative importance of each layer. To further test the robustness of our results, we have performed a sensitivity analysis on these weights. In particular, we have modified their values so that the fraction of secondary infections produced up to the declaration of the National Emergency matches the one typically expected for influenza. Namely, 18% in schools, 19% in workplaces, 30% in households and 33% in the community[23]. Note, however, that the age-susceptibility for influenza is quite different than for COVID-19, and thus it is highly unlikely that this is the real distribution of secondary infections.

Using this approach, we find that we can match the distribution of secondary infections previously described by simply reducing the contribution of workplaces. In particular, we have to reduce the weight of the workplace layer by 70%. In figure S20, we show that this does not alter the overall trend in the community layer, although the lower contribution of the workplace layer produces an increase in the total number of infections in the community. As such, even though the total contribution of each specific layer might change due to the weight distribution, we observe that the general dynamics described in the manuscript are robust.

##### 7.10 Comparison with simpler models

In this last section, we explore the advantages and disadvantages of using simpler versions of the model. In general, with much simpler models it is straightforward to fit macroscopic characteristics of an outbreak such as the evolution of the number of deaths. In this context, one simply needs to add a coefficient next to the transmissibility parameter that will diminish it when the multiple non-pharmaceutical interventions are in place, and fit it to the observed number of deaths.

For this study much more resolution is required, since this allows us to: (i) leave as the only free parameter the values of $R_0$ and the initial number of infected individuals, without artificially modifying transmissibility at any point (since the reduction will already be contained in the behavior of individuals); (ii) explore the dynamics at the level of individuals, being able to explore how some super-spreading events might unfold and compare it with the estimated values with great precision; and (iii) study the dynamical behavior of certain places, rather than assuming that they are always risky or safe regardless of the behavior of individuals. Admittedly, this last part of the study is the hardest one to compare with real data, but that is because measuring this kind of phenomena is incredibly hard in the field, specially after the very initial phase of an outbreak. This is precisely why this model in particular, an modelling in general, can help to shed some light into the dynamics that cannot be easily captured trough traditional epidemiology. Furthermore, this modeling approach goes beyond the particular case

| Probability of a super-spreading event (%) | | | | |
|---|---|---|---|---|
| Category | New York | | Seattle | |
| | Before 03/13 | After 03/13 | Before 03/13 | After 03/13 |
| Arts/Museum | 7.30 [7.01-7.61] | 0.52 [0.48-0.57] | 0.31 [0.08-0.59] | 0.00 [0.00-0.00] |
| Entertainment | 2.42 [2.35-2.49] | 0.14 [0.13-0.15] | 2.16 [1.72-2.63] | 0.21 [0.06-0.39] |
| Exercise | 1.96 [1.91-2.03] | 0.34 [0.32-0.36] | 1.14 [0.88-1.43] | 0.77 [0.51-1.06] |
| Food/Beverage | 0.53 [0.51-0.55] | 0.17 [0.17-0.18] | 0.17 [0.11-0.23] | 0.13 [0.10-0.17] |
| Grocery | 2.18 [2.12-2.24] | 1.31 [1.30-1.33] | 0.58 [0.37-0.81] | 0.93 [0.85-1.02] |
| Health | 0.14 [0.12-0.16] | 0.11 [0.11-0.12] | 0.00 [0.00-0.00] | 0.06 [0.02-0.10] |
| Other | 1.61 [1.54-1.67] | 0.10 [0.09-0.10] | 0.40 [0.21-0.62] | 0.04 [0.00-0.12] |
| Outdoors | 0.03 [0.01-0.06] | 0.00 [0.00-0.01] | 0.00 [0.00-0.00] | 0.00 [0.00-0.00] |
| Service | 0.59 [0.56-0.62] | 0.18 [0.17-0.18] | 0.01 [0.00-0.02] | 0.10 [0.07-0.13] |
| Shopping | 1.43 [1.39-1.47] | 0.84 [0.83-0.85] | 0.14 [0.05-0.27] | 0.09 [0.06-0.11] |
| Sports/Events | 8.73 [8.32-9.14] | 4.27 [3.90-4.66] | 0.22 [0.00-0.56] | 0.00 [0.00-0.00] |
| Transportation | 0.26 [0.21-0.31] | 0.04 [0.03-0.05] | 0.00 [0.00-0.00] | 0.00 [0.00-0.00] |
| All | 1.73 [1.71-1.75] | 0.71 [0.70-0.71] | 0.93 [0.84-1.02] | 0.34 [0.32-0.37] |

**Table S2.** Probability that an individual will cause a super-spreading event as defined in[20]. We aggregate all the infections produced by each individual within each category for the given period of time, and compute the fraction of individuals who produce a super-spreading event out of the total number of individuals infecting someone in that category. In brackets the 95% C.I. computed using a bootstrap percentile method is shown.

of SARS-CoV-2 and could be applied to future pandemics. Understanding that the role that some places might play in the propagation of an emergent disease is a dynamic process, which depends not only on the characteristics of the place but also on the behavior of individuals is thus of paramount importance.

To demonstrate the information that can be obtained using different levels of resolution, we explore three simplified versions of our model:

1. Substituting the community layer by a network in which all the nodes who have been observed at least once in the community are present every day, but connections are established at random each day with an average degree $\langle k \rangle = 10$. Since the results could depend on the choice of $\langle k \rangle$, we further weight the links so that the average strength $\langle s \rangle$ (sum of the weights of each node) is equal to the one contained in the full model. This also adds the effect of the reduction in the strength observed in the data during the lockdown phase.

2. As the previous model, but with $\langle k \rangle = 20$.

3. As in 2), but every day only the nodes observed in the real data are present. Thus, this model can be described as a complete randomization of the connections each day (so that the information on where those connection happened is lost) and with $\langle k \rangle = 20$.

In figure S21, we show the results of the calibration process in these networks. The corresponding AIC for each model is: 601.59 for the one using real data; 5692.04 for the one with fixed $N$ and $\langle k \rangle = 10$; 5773.02 for the one with fixed $N$ and $\langle k \rangle = 20$; and 647.58 for the one with variable number of nodes and $\langle k \rangle = 20$. Between the best two models, the $\Delta AIC = 45.99$, yielding a relative likelihood of Rel. Like. $= e^{-\frac{1}{2}\Delta AIC} = 10^{-10}$. As such, of these four models, the one that betters fits the data is the original one described in the manuscript. Several things are worth highlighting:

1. The two models that incorporate all the nodes in the community layer cannot be fitted properly to the data. In essence, if $R_0$ is too low the spreading is too slow. Conversely, for larger values the speed is correct, but the total amount of deaths is much higher. The results shown in the picture where obtained after increasing the MAE used in the calibration of the main model from 25 to 80, since no runs entered within the original threshold.

2. The result does not depend on the value of $\langle k \rangle$. This is to be expected since the average strength of each node, $\langle s \rangle$, is the same in both cases.

3. When both the average strength (so that the specific choice of $\langle k \rangle$ does not play a role) and the diminishing of the total number of nodes present in the community layer (as a consequence of the non-pharmaceutical interventions and the stay-at-home mandates) are taken into account, it is possible to fit the model as in the complete scenario.

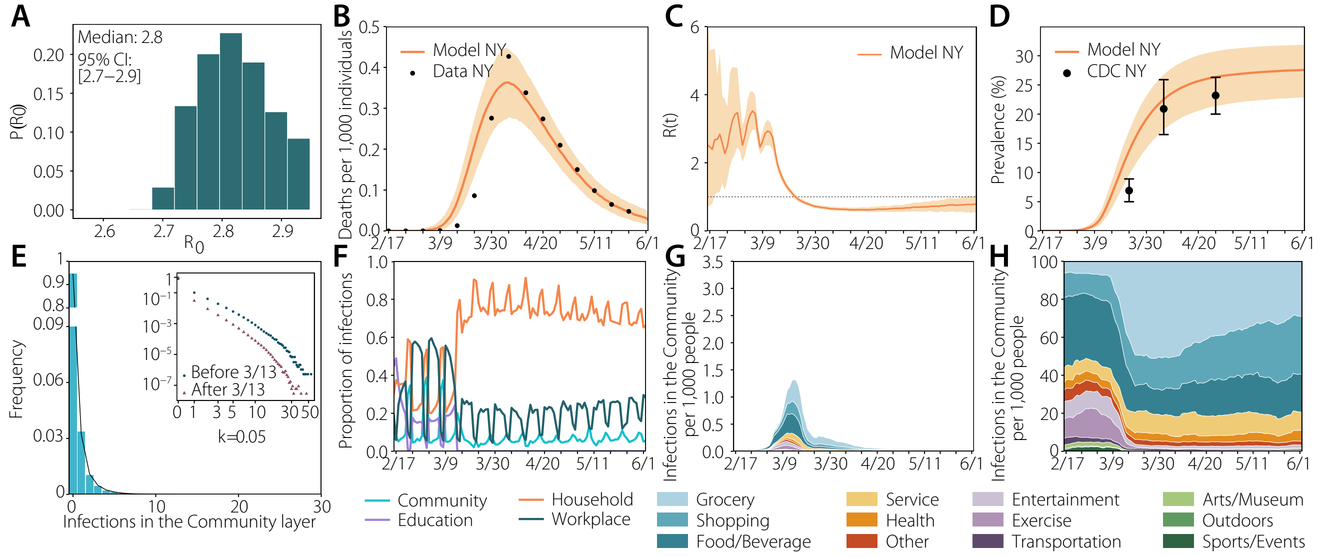

**Figure S10.** Results with a more restricted definition of stay for the case of New York (10 meters): (a) estimated $R_0$; (b) number of deaths (fit); (c) estimated $R_t$; (d) prevalence; (e) distribution of infections; (f) proportion of infections per layer; (g) infections per setting; (h) normalized infections per setting.

The main problem behind models (1) and (2) is that, even though the strength is correctly adjusted, the presence of many more individuals in the community than what actually happened increases too much the transmissibility. To solve this, some models propose to reduce the transmissibility parameter ($\beta$ in our case) by an amount extracted from the fitting. Another possibility would be to compute the fraction of individuals that is observed each day and use it to reduce the transmissibility, but that would imply using almost the same information that the full model contains, not making this choice any simpler.

Thus, in order to properly model the evolution of the outbreak in New York without adding more assumptions than the bare minimum that are needed to work with the real data, at the very least we need to take into account the number of individuals observed each day and the average strength (so that the choice of $\langle k \rangle$ will not play a role). This simplified version of the model, even though able to capture the overall evolution of the outbreak, has nonetheless some limitations.

First, if we look at the distribution of the number of secondary infections (Fig. S22) we observe that this model does not yield large super-spreading events. The reason is that the choice of the degree distribution has a direct impact on these events. To obtain a super-spreading event it is necessary to have some heterogeneity, either in the transmissibility of specific settings (for which we need information of where the individuals where plus an estimation of how the characteristics of each particular place affect the spreading, something that even nowadays are still unknown), or in the number of connections of each node. Our choice of degree distribution in the simplified versions of the model is the homogeneous distribution or random network model. Of course, it would be possible to choose any other distribution, but that would imply making further assumptions on the type of the distribution and on its parametrization. With the proposed approach, however, the heterogeneity comes directly from the observed behavior of individuals.

Second, even if one uses an heterogeneous degree distribution while building the simplified community layer, the information of the settings where infections may have taken place would be lost. Indeed, if we want to be able to say something about the role of specific settings in the propagation of the first wave in New York, it is thus necessary to resort to a model with this level of detail.

To summarize, the modeling approach we proposed in this manuscript is the simplest one of the four that can: (i) fit the evolution with minimal assumptions and without adding any external events not contained in the data; (ii) describe the distribution of the secondary number of infections; and (iii) shed some light on the dynamical role that some settings may play in the context of a pandemic and their relationship with the behavior of individuals.

Furthermore, going to the individual level allows us to measure things that other models that use aggregated data cannot. For instance, we can explore the number of secondary infections per individual, and observe the presence of over-dispersion. The model that uses real data is the one that best matches the estimates found in the literature[24].

This also allows us to explore the dynamic behavior of each setting in terms of potential super-spreading events. Admittedly, the characteristics of certain locations might yield them more prone to such events, but the specific details of this are still unknown.

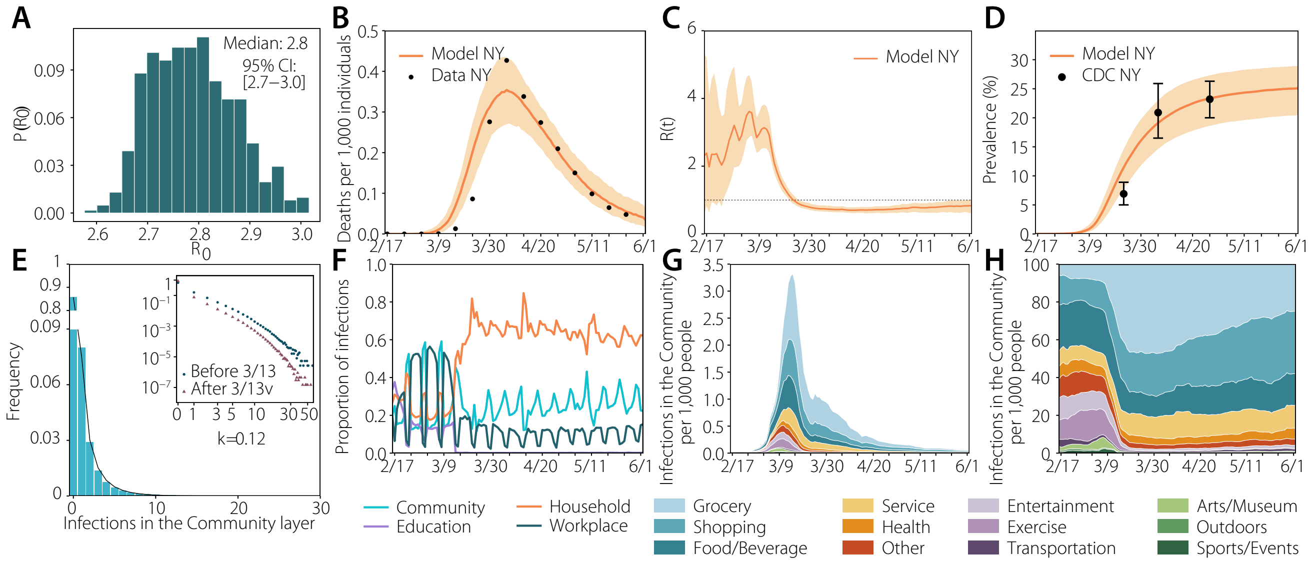

**Figure S11.** Main results in New York with larger pre-symptomatic transmissibility: (a) estimated $R_0$; (b) number of deaths (fit); (c) estimated $R_t$; (d) prevalence; (e) distribution of infections; (f) proportion of infections per layer; (g) infections per setting; (h) normalized infections per setting.

| Model | Over-dispersion |
|---|---|
| Real data | 0.16 |
| Fixed N $\langle k \rangle = 10$ | 3.02 |
| Fixed N $\langle k \rangle = 20$ | 3.01 |
| Var. N $\langle k \rangle = 10$ | 0.34 |

**Table S3.** Over-dispersion in the number of secondary infections obtained in each of the four models considered

Another quantity that can be measured in our model that it is not available in more aggregated models is the household secondary attack rate. That is, the attack rate of a household in which the index case in said household is not taken into account. In figure S23, we show how this quantity evolves with time. In the early phase of the pandemic, it is close to 20%, in line with what is reported in the literature[25]. Then, during the stay-at-home period, this quantity is greatly increased reaching very high values. This evolution might change depending on the assumptions behind the model, such as whether an infected individual will try to self-isolate from the rest of the family or not. This will further depend on the help provided by the government on this regard, the size of the households, etc. Although we did not focus on these elements for this study, they could be added in the future to improve the model.

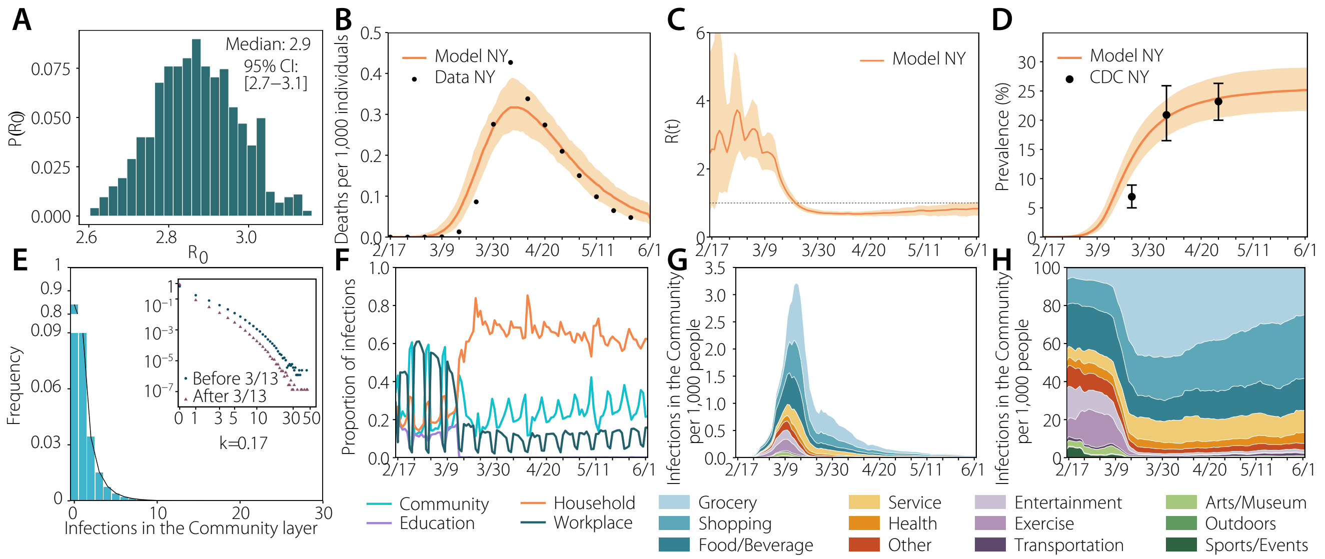

**Figure S12.** Main results in New York with longer time to death notification: (a) estimated $R_0$; (b) number of deaths (fit); (c) estimated $R_t$; (d) prevalence; (e) distribution of infections; (f) proportion of infections per layer; (g) infections per setting; (h) normalized infections per setting.

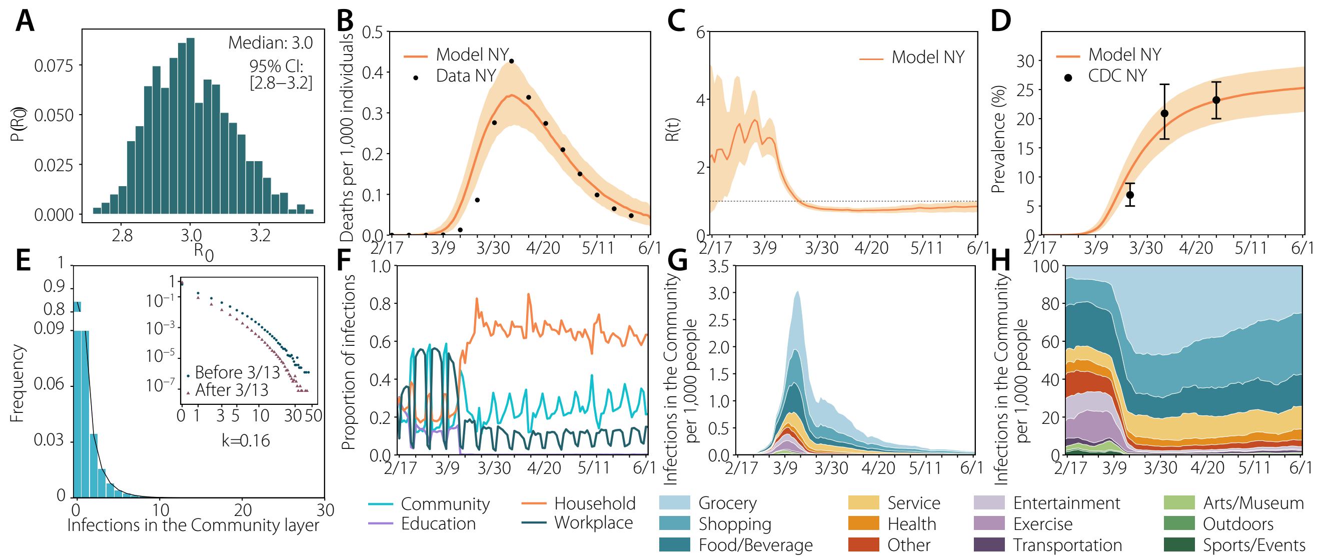

**Figure S13.** Main results in New York with larger outdoor transmissibility: (a) estimated $R_0$; (b) number of deaths (fit); (c) estimated $R_t$; (d) prevalence; (e) distribution of infections; (f) proportion of infections per layer; (g) infections per setting; (h) normalized infections per setting.

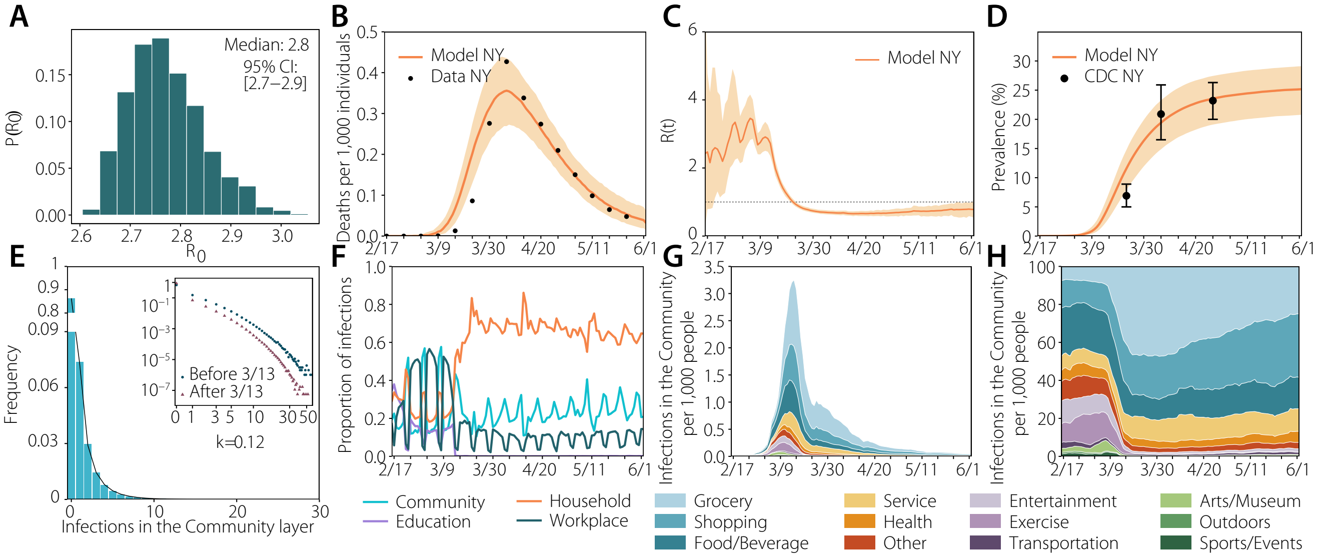

**Figure S14.** Main results in New York without symptomatic transmission: (a) estimated $R_0$; (b) number of deaths (fit); (c) estimated $R_t$; (d) prevalence; (e) distribution of infections; (f) proportion of infections per layer; (g) infections per setting; (h) normalized infections per setting.

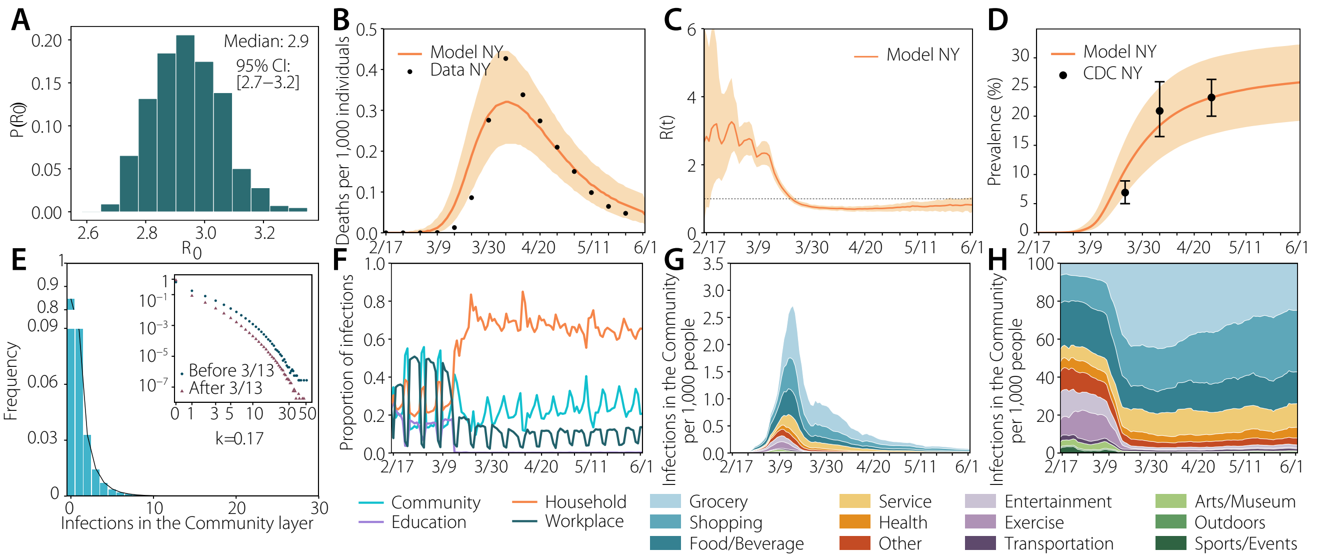

**Figure S15.** Results with a resampled population to remove economic bias in New York: (a) estimated $R_0$; (b) number of deaths (fit); (c) estimated $R_t$; (d) prevalence; (e) distribution of infections; (f) proportion of infections per layer; (g) infections per setting; (h) normalized infections per setting.

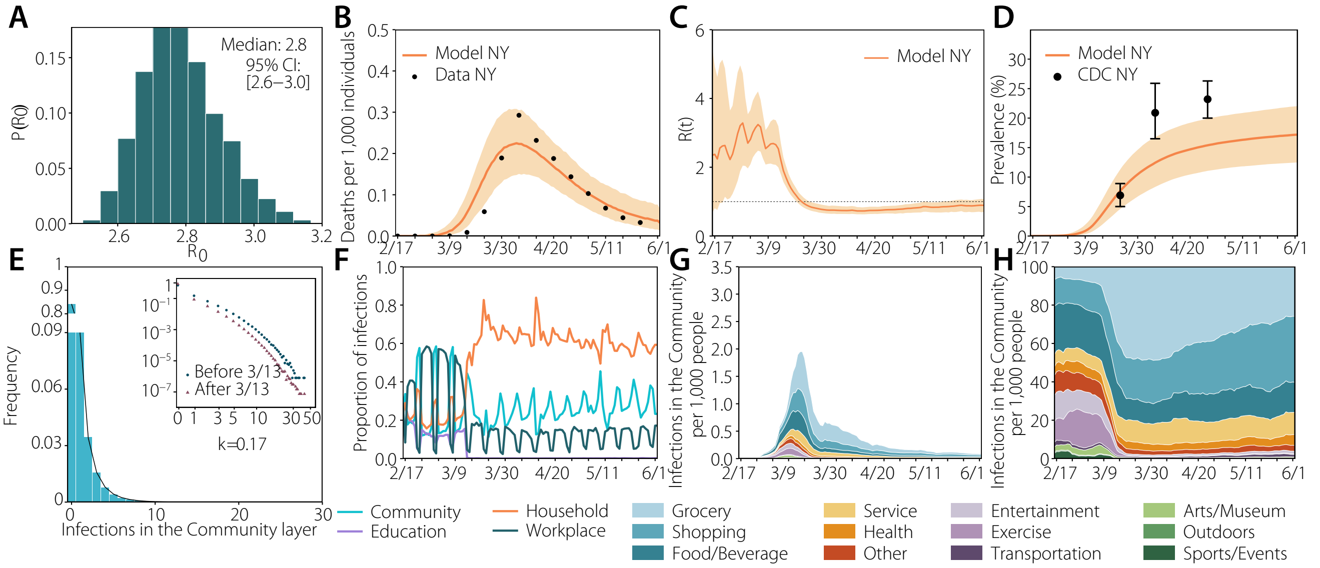

**Figure S16.** Results when the model is fitted to a smaller number of deaths: (a) estimated $R_0$; (b) number of deaths (fit); (c) estimated $R_t$; (d) prevalence; (e) distribution of infections; (f) proportion of infections per layer; (g) infections per setting; (h) normalized infections per setting.

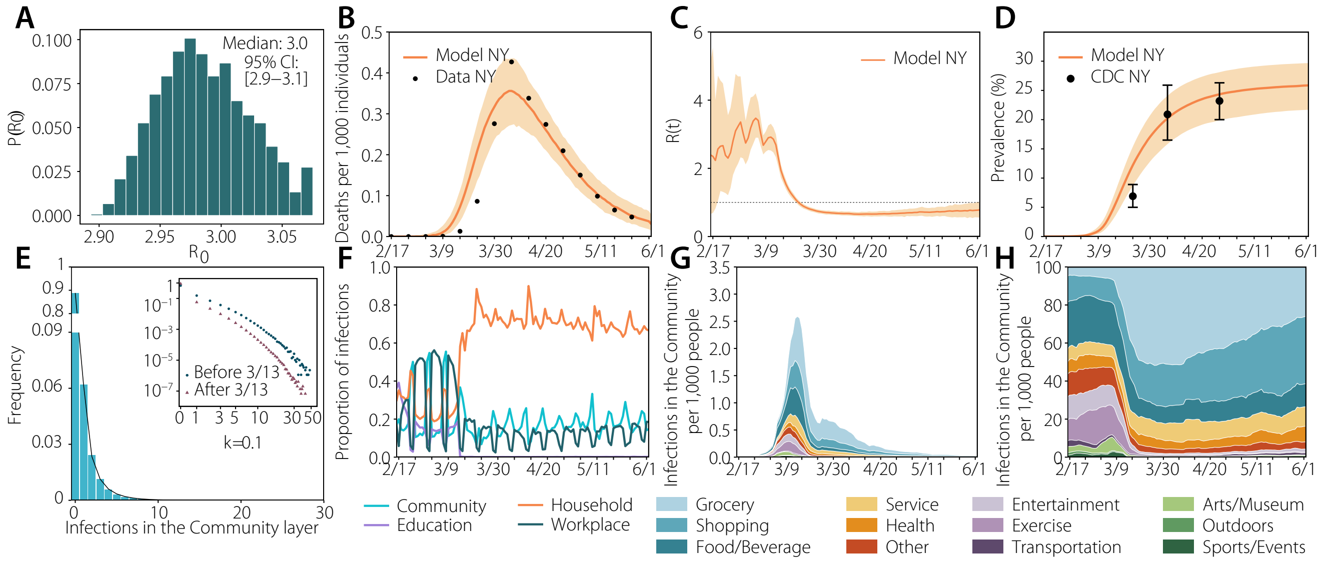

**Figure S17.** Results with stricter definition of stay in New York (minimum 15 minutes): (a) estimated $R_0$; (b) number of deaths (fit); (c) estimated $R_t$; (d) prevalence; (e) distribution of infections; (f) proportion of infections per layer; (g) infections per setting; (h) normalized infections per setting.

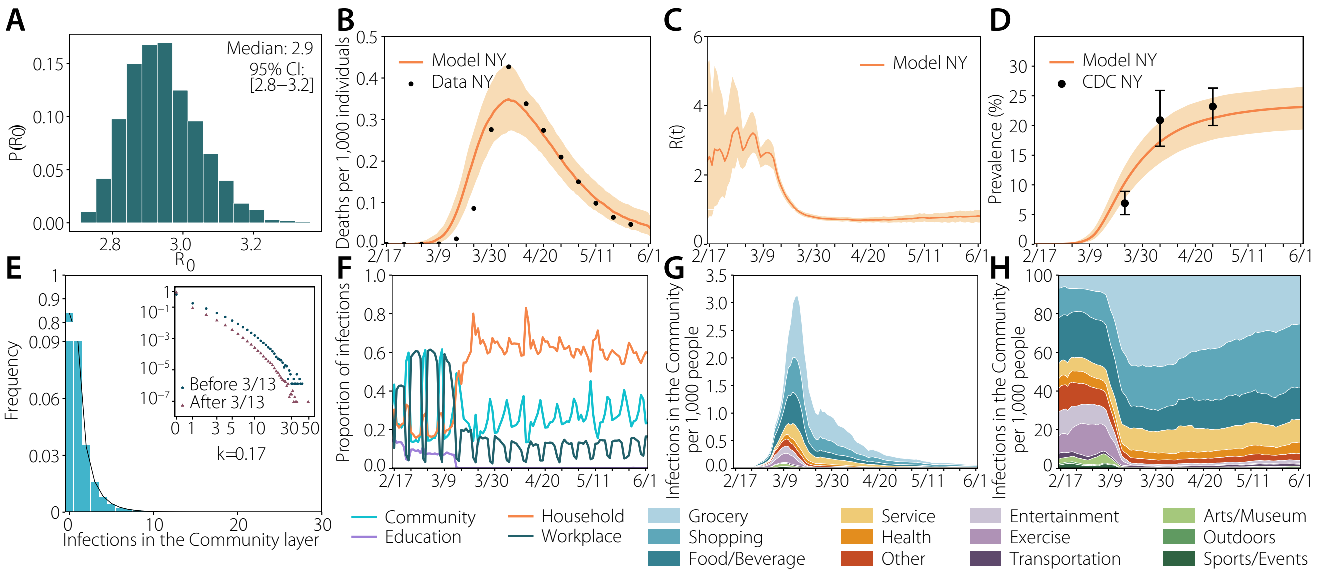

**Figure S18.** Results with differential age-susceptibility: (a) estimated $R_0$; (b) number of deaths (fit); (c) estimated $R_t$; (d) prevalence; (e) distribution of infections; (f) proportion of infections per layer; (g) infections per setting; (h) normalized infections per setting.

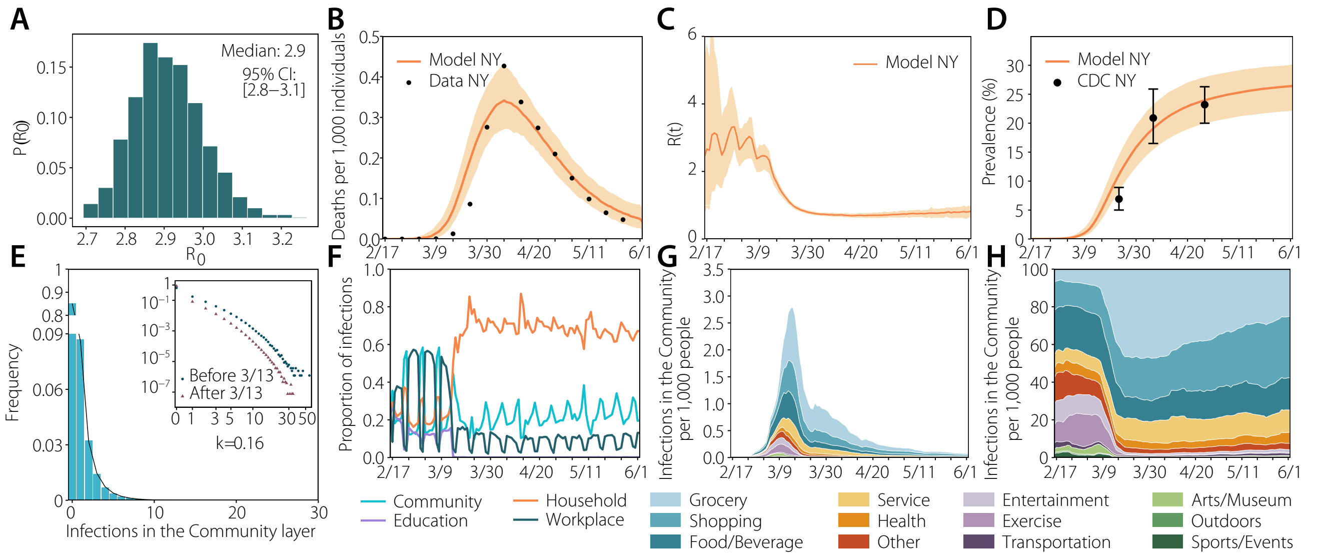

**Figure S19.** Results with larger household transmissibility after the declaration of the N.E.: (a) estimated $R_0$; (b) number of deaths (fit); (c) estimated $R_t$; (d) prevalence; (e) distribution of infections; (f) proportion of infections per layer; (g) infections per setting; (h) normalized infections per setting.

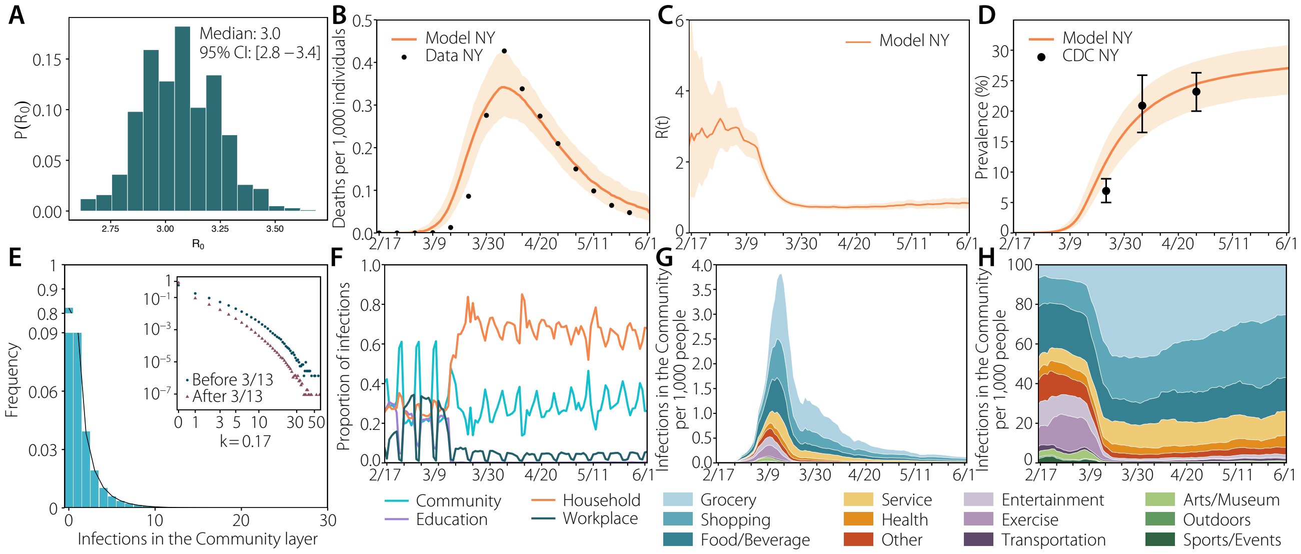

**Figure S20.** Results with layer's weight calibrated to the distribution of secondary infections per setting of influenza (18% in schools, 19% in workplaces, 30% in households and 33% in the community): (a) estimated $R_0$; (b) number of deaths (fit); (c) estimated $R_t$; (d) prevalence; (e) distribution of infections; (f) proportion of infections per layer; (g) infections per setting; (h) normalized infections per setting.

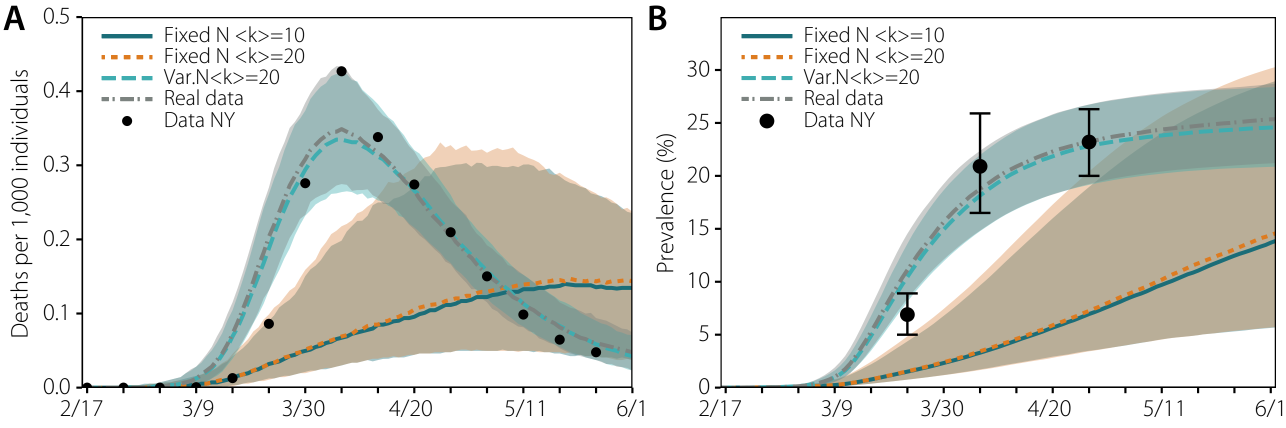

**Figure S21.** Fitting the four models proposed. a) Evolution of the number of deaths as a function of time for each model, fitted to the real data. b) Prevalence extracted from the output of the model (not fitted).

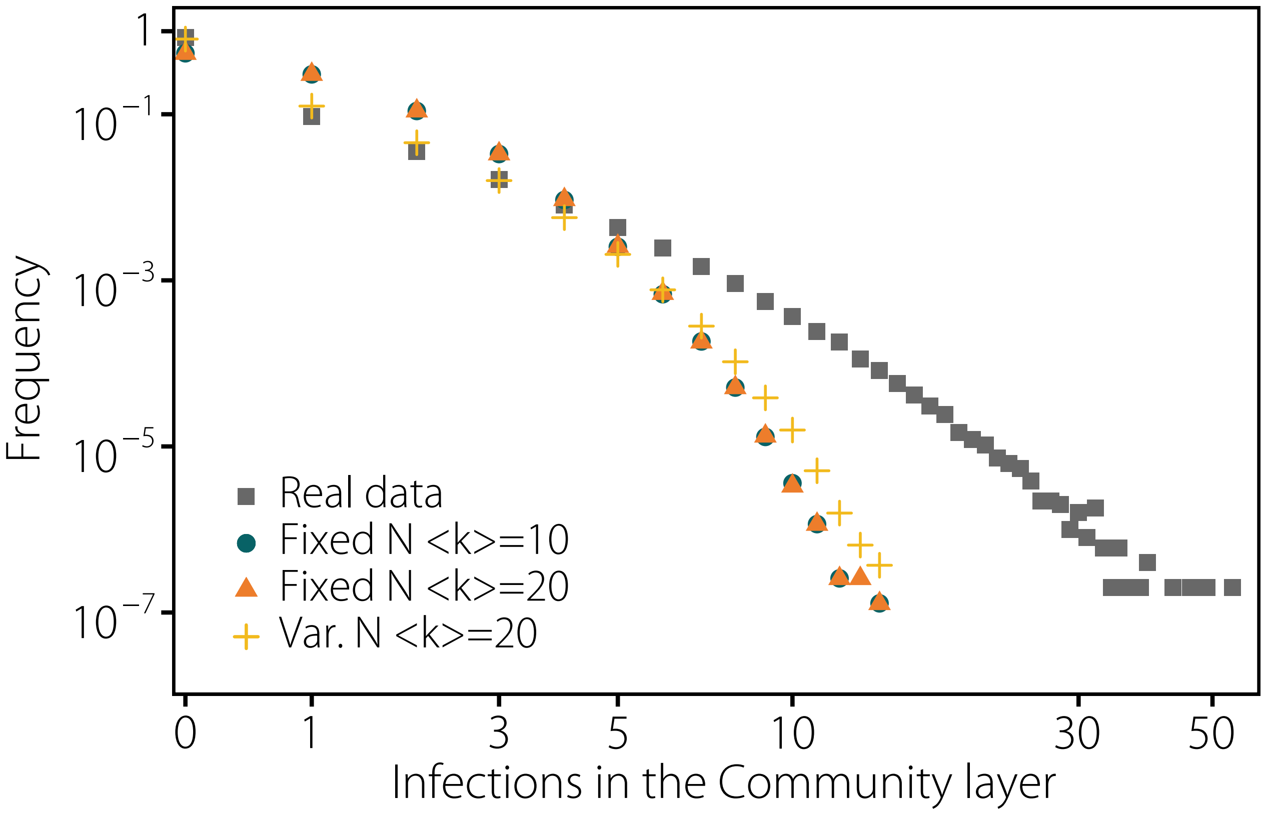

**Figure S22.** Distribution of the number of secondary infections in each of the models considered. Only the model with heterogeneous degree distribution yields a distribution of the number of secondary infections compatible with super-spreading events.

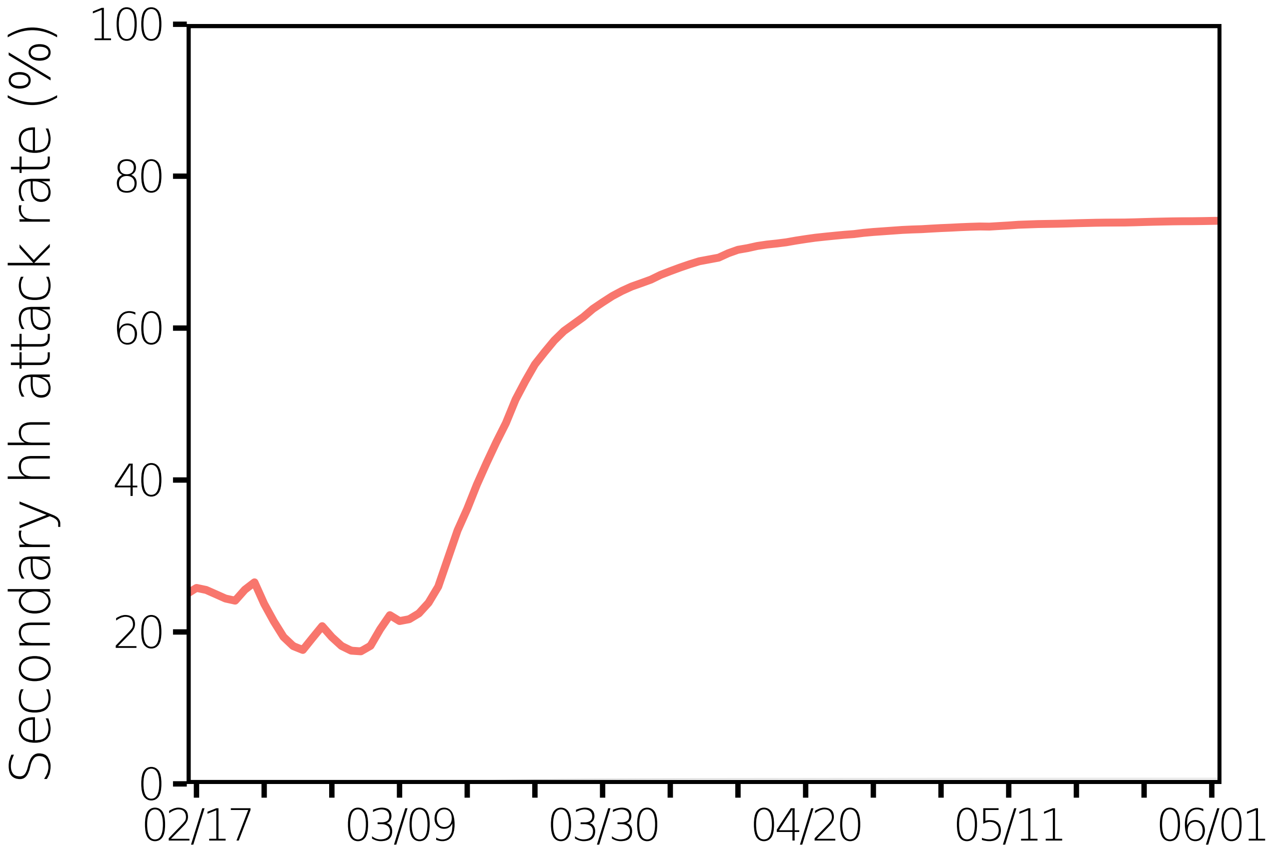

**Figure S23.** Household secondary attack rate in the baseline scenario for NY described in the main paper.
